# An SEIARD epidemic model for COVID-19 in Mexico: mathematical analysis and state-level forecast

**DOI:** 10.1101/2020.05.11.20098517

**Authors:** Ugo Avila-Ponce de León, Ángel G. C. Pérez, Eric Avila-Vales

## Abstract

We propose an SEIARD mathematical model to investigate the current outbreak of coronavirus disease (COVID-19) in Mexico. Our model incorporates the asymptomatic infected individuals, who represent the majority of the infected population (with symptoms or not) and could play an important role in spreading the virus without any knowledge. We calculate the basic reproduction number (*R*_0_) via the next-generation matrix method and estimate the per day infection, death and recovery rates. The local stability of the disease free equilibrium is established in terms of *R*_0_. A sensibility analysis is performed to determine the relative importance of the model parameters to the disease transmission. We calibrate the parameters of the SEIARD model to the reported number of infected cases and fatalities for several states in Mexico by minimizing the sum of squared errors and attempt to forecast the evolution of the outbreak until August 2020.

## 1 Introduction

At the beginning of December 2019, a new virus caused an increase in atypical pneumonia at the city of Wuhan in China. The virus was isolated, sequenced and identified as a new type of coronavirus [1]. The virus was called SARS-CoV-2 and the disease associated with that virus was called COVID-19 [2]. On March 11, 2020 the World Health Organization (WHO) declared the outbreak of COVID-19 as a global pandemic [3]. Most of the infected individuals will develop a mild respiratory illness and they won’t need any special requirements; they will just need to manage the symptoms. Symptoms associated to COVID-19 are fever, cough and fatigue, few may develop other symptoms like headache and diarrhea to name some.

The first case of COVID-19 confirmed in Mexico was until February 2020, which was an imported case from Europe, by that time it was an epicenter of the disease. Mexican authorities announced on March 14th the “Jornada Nacional de Distanciamiento Social” that basically means quarantines for vulnerable groups. Not only elderly individuals are considered a group of risk, individuals that have comorbidities like diabetes, hypertension and obesity can develop more complicated respiratory symptoms that can be fatal. Mexico declared phase 2 of the coronavirus pandemic on March 23 with 367 confirmed cases. As of May 1st, there were 20 739 confirmed cases and 1972 deaths. In [4], Cruz-Pacheco et al. estimated the arrival of the infectious outbreak to Mexico between March 20 and March 30, 2020. Other models for predicting the evolution of COVID-19 outbreak in Mexico have been proposed in [5, 6].

Compartmental models like the one used in this paper have been used for studying the spread of the COVID-19 pandemic in several countries, such as China [7, 8] and Italy [9, 10]. In this study, we use a compartmental mathematical model to try to understand the outbreak of COVID-19 in Mexico and we evaluate the heterogeneity of COVID-19 throughout Mexican territory considering two important regions: the Mexico Valley and the Yucatan Peninsula.

The rest of the paper is structured as follows. We formulate the mathematical model, compute the basic reproduction number and perform a sensibility analysis in Section 2. In Section 3, we calibrate our mathematical model using a sum of squared errors approach using daily cumulative of infected and death individuals published daily by the Ministry of Health in Mexico, we used the data until May 4th. In Section 4, we explore the simulations of the cases in Mexico and we compare how different the outbreak is in the two regions. Lastly, we provide some concluding remarks in Section 5.

## 2 Mathematical model with asymptomatic individuals

In this work, we will use a compartmental differential equation model for the spread of COVID-19 in Mexico. The model monitors the dynamics of six subpopulations, which are: susceptible (*S*(*t*)), exposed (*E*(*t*)), infected (*I*(*t*)), asymptomatic (*A*(*t*)), recovered (*R*(*t*)) and dead (*D*(*t*)).

The model simulations will be carried out with the following assumptions:

a. Individuals of 12 years old and higher are susceptible to the virus.
b. The susceptible and infected individuals are homogeneous in the population.
c. At first, no interventions were applied to stop the spread of COVID-19.
d. The population is constant; no births are allowed, and we only take into account the fatalities associated to COVID-19.

Figure 1 shows a diagram of the flow through the compartmental subpopulations.

**Figure 1:**
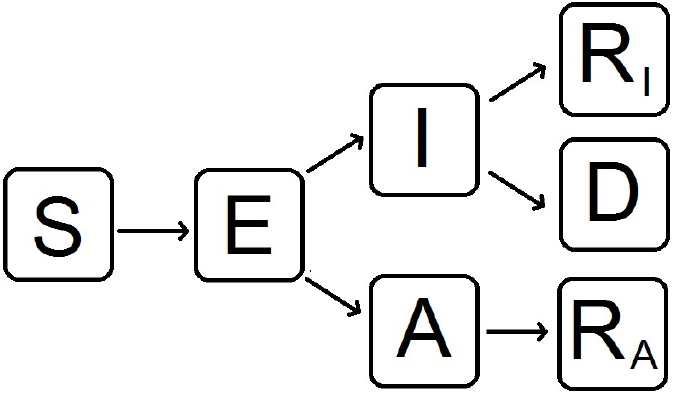
Flow diagram of our mathematical model to evaluate the behavior of the spread of nCoV-2019 in Mexico. *S*: susceptible, *E*: exposed, *I*: infected, *A*: infected but without symptoms (asymptomatic), *R_I_*: recovered from symptomatic infection, *R_A_*: recovered from asymptomatic infection, *D:* dead.

**Susceptible population** *S*(*t*): This subpopulation will remain constant because recruiting individuals is not allowed in our model. The susceptible population will decrease after an infection, an acquired characteristic due to the interaction with an infected person or asymptomatic one. The transmission coefficients will be *βI* and *βA*. The rate of change of the susceptible population is expressed in the following equation:

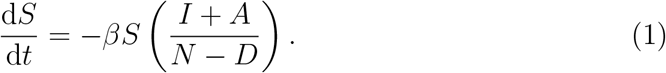

**Exposed population** *E*(*t*): This subpopulation consists of individuals that are infected but cannot infect others. The population decreases at a rate *w* to become infected or asymptomatic. Consequently,

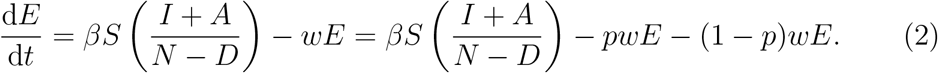

**Infected population** *I*(*t*): Infected (symptomatic) individuals are generated at a proportion *p* (*p* ∊ (0,1)) from the exposed class. They recover at a rate *γ* and die at a rate *δ*. This is the only population that acknowledges death. Thus,

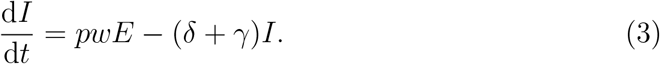

**Asymptomatic population** *A*(*t*): This population is considered an infected population, but the individuals do not develop the common symptoms of COVID-19. Asymptomatic individuals are important to model because they have the ability to spread the virus without knowing; they are produced at a rate 1 − *p* and recover at a rate *γ*. Consequently,

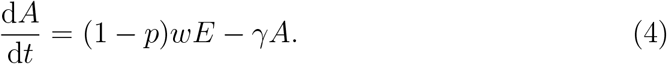

**Recovered populations** *R_I_*(*t*) **and** *R_A_*(*t*): All individuals infected with symptoms or not will recover at a rate *γ*. We subdivide the recovered population in two compartments: individuals who recover after having symptoms (*R_I_*) and individuals who recover from asymptomatic infection (*R_A_*). Hence

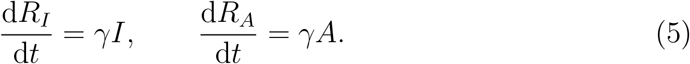

**Dead population** *D*(*t*): Infected individuals with symptoms die at a rate *δ*, that is,

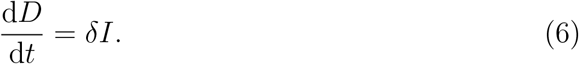

Hence, the system of differential equations that will model the dynamics of coronavirus spread in Mexico is:

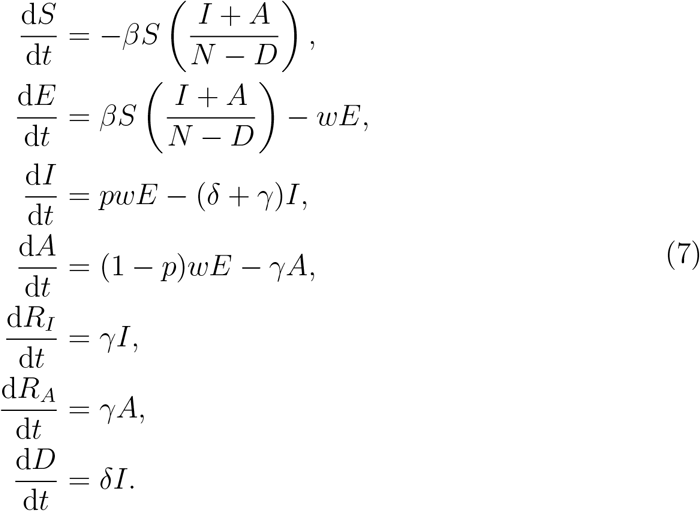

We also observe that *N*:= *S* + *E* + *I* + *A* + *R_I_* + *R_A_* + *D* is constant, where *N* is the size of the population modeled.

### 2.1 Basic reproduction number with a disease-free equilibrium

There exists a disease-free equilibrium, which is given by *S* = *N*, *E* = *I* = *A* = *R_I_* = *R_A_* = *D* = 0, and we will denote it by *x*_0_. We calculate the basic reproduction number *R*_0_ based on this steady state. By applying the next-generation matrix method to find *R*_0_, we must solve the equation *R*_0_ = *ρ*(*FV^−^*^1^), where *F* and *V* are the derivatives of the new infections matrix 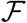 and the transition matrix 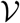, respectively, evaluated at the disease-free equilibrium, and *ρ* denotes the spectral radius. Then

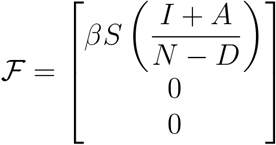

The derivative of 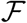 at *x*_0_ is:

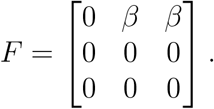

The transition matrix is

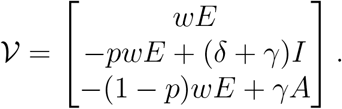

The derivative of 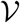 at *x*_0_ is

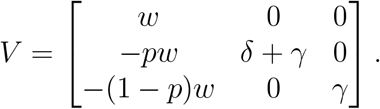

The inverse of *V* is

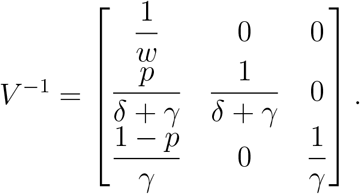

Then

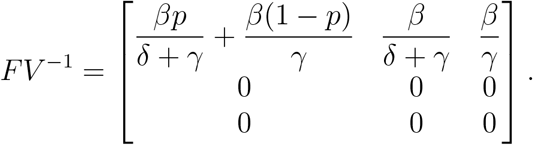

We need to find the eigenvalues of *FV*^−1^, which are 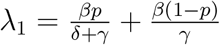, *λ*_2_ = 0 and *λ*_3_ = 0. Then, the basic reproduction number is given by the dominant eigenvalue, that is,

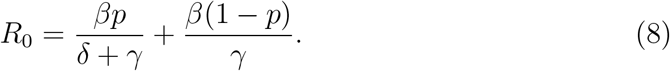

The basic reproduction number formulated above has two components that have a biological interpretation. The first half of (8) is associated with symptomatic individuals: during the time of recovery 1/*γ* they can infect individuals at a rate *β*, only these infected individuals die at a rate 1/*δ*. Thus, the first term of (8) can be re-written as:

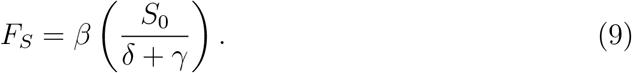

The same derivation is for the second half of (8), only this represents the asymptomatic individuals. Hence,

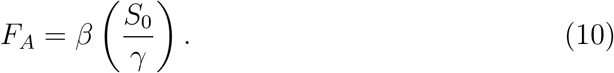

The functionality of the basic reproduction number is associated with the force of infection of both symptomatic and asymptomatic individuals. Thus, (8) can be expressed by the following equation:

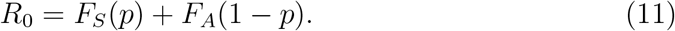

The stability of our disease-free equilibrium can be explained by the following theorem, mentioned in [11]: the disease-free equilibrium *NC* = (*N*, 0, 0, 0,0, 0, 0) of our system of differential equations is locally asymptomatically stable if *R*_0_ < 1 and unstable if *R*_0_ > 1.

We calculate the Jacobian Matrix of our system of differential equations at the disease-free equilibrium, which is given by

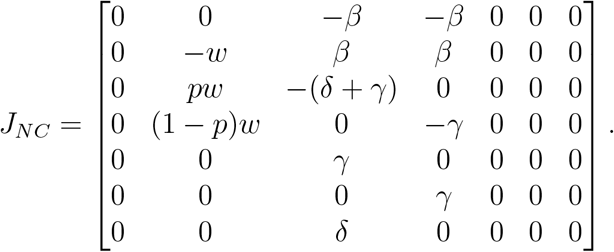

Let λ be the eigenvalue of the matrix *J_NC_*. Then the characteristic equation is given by det(*J_NC_* − *λI*) = 0, that is,

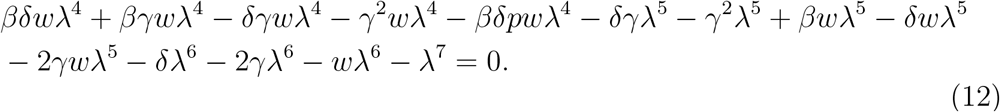

To solve (12), we can factorize with *λ*^4^. Thus, (12) can be re-written as

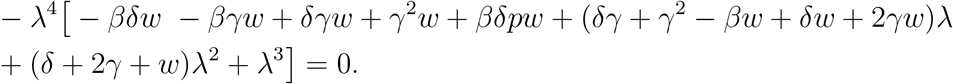

To determine the stability of the disease-free equilibrium, we must solve the cubic equation

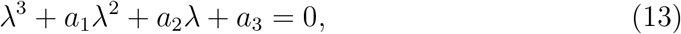

where the coefficients are

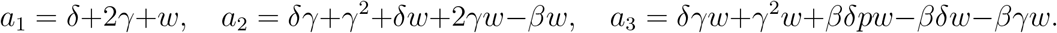

The Routh-Hurwitz criterion tells us that a necessary and sufficient condition for all roots of (13) to have negative real part is that *a*_3_ > 0 and *a*_1_*a*_2_ − *a*_3_ > 0. In order to check these conditions, we rewrite *a*_3_ in terms of the basic reproduction number as *a*_3_ = *γw*(*δ* + *γ*)(1 − *R*_0_). From this, it is clear that *a*_3_ is positive whenever *R*_0_ < 1. Therefore, we can conclude the following result:

#### Theorem 1

The disease-free equilibrium of model (7) is stable if and only if *R*_0_ < 1 and

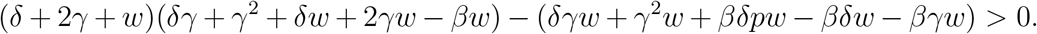

### 2.2 Sensitivity analysis of the basic reproduction number

Using the formula (8) for the basic reproduction number, we can perform a sensitivity analysis for *R*_0_ to determine how important each parameter is for disease transmission. Sensitivity indices allow us to measure the relative change in a variable, in this case *R*_0_, when a parameter varies. This can be used to determine the robustness of model predictions to parameter values, and to discover parameters that have a very high impact on *R*_0_ [12]. Hence, we use the following definition.

#### Definition 1

If *R*_0_ is differentiable with respect to a given parameter *θ*, the normalized forward sensitivity index of *R*_0_ is defined by

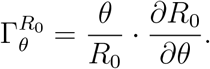

We will calculate the sensitivity index for *R*_0_ with respect to the parameters *β*, *p*, *δ* and *γ* as follows (notice that *R*_0_ does not depend on *w*, so 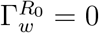).

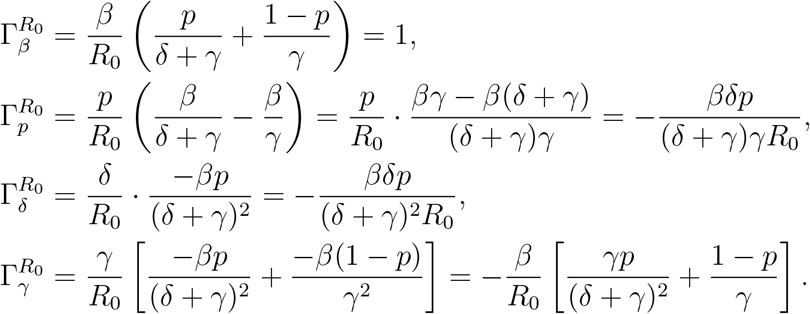

From this, we can see that 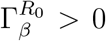, while 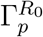, 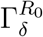, 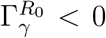, which means that an increment in the contact rate *β* will cause *R*_0_ to increase, while an increment in the symptomatic case proportion *p*, the death rate *δ* or the recovery rate *γ* will cause *R*_0_ to decrease.

Also, we can see that

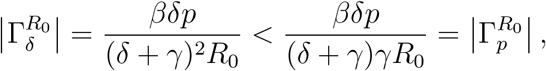

which implies that a perturbation in the parameter p produces a relatively larger change in *R*_0_ than a perturbation in *δ*.

Since 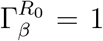, we can see that increasing the contact rate *β* by a given percentage always increases *R*_0_ by that same percentage. Moreover, 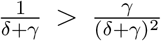, so

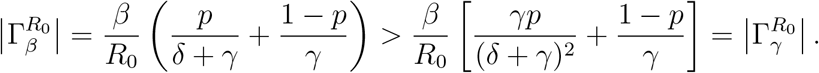

Hence, the parameters with the largest normalized forward sensitivity index are *β* and *p*.

## 3 Implementation to estimate the parameters

To describe the evolution of the epidemic in Mexico taking into account the social distancing measures taken by the government, we will assume that the infection rate, recovery rate and death rate are time-dependent functions, similar to those used in [10].

To model the effect of epidemic control measures, which cause the number of contacts per person per unit time to decrease as the epidemic progresses, we describe the infection rate by the function

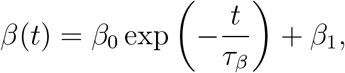

where *β*_0_ + *β*_1_ is the initial infection rate. This rate decreases exponentially to the value *β*_1_ with a characteristic time of decrease *τ_β_*.

The time of recovery for patients may also vary with time due to the medical staff improving their therapeutic procedures. Hence, we will assume that the recovery rate is modeled by the function

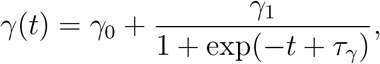

where *γ*_0_ is the recovery rate at time zero, and γ_0_ + γ_1_ is the recovery rate at a later time, which is reached after τ_γ_ days of adaptation.

Lastly, the death rate may decrease with time due to the adaptation of the pathogen or the development of more advanced treatments. Hence, we can model this with the function

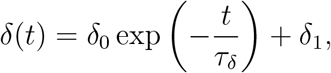

where *δ*_0_ + *δ*_1_ is the initial death rate, which decreases to the value *δ*_1_ with a characteristic time *τ_δ_*.

If we replace the constant parameters *β*, *δ* and *γ* in equation (8) with the aforementioned time-dependent functions, we can define

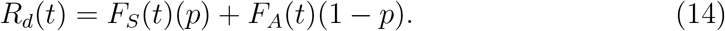

As we derived before, *F_S_* and *F_A_* are represented by *β*, *δ* and *γ*. *R_d_*(*t*) is the effective daily reproduction ratio, which measures the number of new infections produced by a single infected individual per day, taking into account the evolving public health interventions and available resources [13].

The set of differential equations was solved using Matlab 2016b with the ode45 solver, which is based on an explicit Runge-Kutta (4,5) formula. Our model was calibrated using the cases of COVID-19 in Mexico. The data were collected in the period since the first day after Mexico declared phase 2 of the pandemic (March 12th) until May 4th from the open source repository of the Ministry of Health in Mexico [14].

The optimization of parameters to describe the outbreak of COVID-19 in Mexico was fitted by minimizing the Sum of Squared Errors (SSE), in such a way that the solutions for *D*(*t*) obtained by the model approximate the reported cumulative number of fatalities, while the sum *I*(*t*) + *R_I_*(*t*) + *D*(*t*) approximates the reported cumulative number of infected cases with symptoms. Since the Mexican government does not keep a record of the number of asymptomatic cases, we assume that the asymptomatic infected population is about nine times larger than the population with symptoms, based on government estimations.

Since we do not have a reliable report of the number of recoveries by state in Mexico, we choose not to fit the values of the recovery-related parameters (*γ*_0_, *γ*_1_ and *τ_γ_*) and optimize only the values of the other parameters (*β*_0_, *β*_1_, *τ_β_*, *δ*_0_, *δ*_1_, *τ_δ_*, *w* and *p*). For parameters *γ*_0_, *γ*_1_ and *τ_γ_* we use the same fixed values for all states, which were obtained previously in a best fit optimization using the recovered data for all Mexico [15].

We applied three searches to minimize the SSE function: a gradient-based method, a gradient-free algorithm, and finally, a gradient-based method. This method was necessary to obtain the global minimum. We adapted the code from Caccavo [10] for our mathematical model. The code and the data will be available in the following github: https://github.com/UgoAvila/COVID-19.Mexico.

## 4 Evolution of the outbreak of COVID-19

### 4.1 Evolution of the outbreak of COVID-19 in mainland Mexico

The predicted evolution of the outbreak for COVID-19 in Mexico can be seen in Figure 2. The parameters of the mathematical model were fitted with the experimental data provided by a daily update from the Mexican Ministry of Health. By adjusting the data from the period from March 12, 2020 to April 28, 2020, we simulated the daily new COVID-19 cases in Mexico until August 2020. The peak of the infection modeled will be in the middle of May, with 13 000 infected individuals with the known developed symptoms and roughly 130 000 infected individuals that will not develop any kind of symptoms. The values of the best fit parameters are given in Table 1. Figure 3 shows the variation of the infection rate *β*(*t*), recovery rate *γ*(*t*) and death rate *δ*(*t*) with respect to time. The infection rate decreases at a good pace with respect to time, taking roughly 90 days from March 12 to a near infection rate per day of 0, which means that from that day they will be no more new infected individuals. The recovery rate in Mexico at twenty days past March 12 is increasing, and past the 20th day the recovery rate becomes constant with respect to time. This type of behavior may be explained by the fact that, at first, there were only mild cases in Mexico. Then, by March 30 there were much more severe cases, which are associated with a death rate that remains roughly constant at a rate of 0.015 per day. Using these values for the parameters, we can calculate the effective daily reproduction ratio *R_d_*(*t*) for each day (see Figure 4) As we can see in Figure 4, during March *R_d_*(*t*) decreases exponentially from 16 to 4, roughly, and it decreases from 4 to 2 during the month of April. The decrease from the first period is very helpful and Mexico will be entering a declared phase two of the pandemic with a relatively low reproduction number. We believe this decay may have occurred because the Ministry of Health applied the “Jornada de Distanciamiento Social” before Mexico declared to be in phase two. Our simulation shows that *R_d_*(*t*) will become less than 1 around the second week of May 2020. In Figures 5–8, we carried out the simulation with the best fit parameters. We show a comparison of the cumulative number of infections (Figure 5) and deaths (Figure 6) with the reported data. We also plot the number of active symptomatic infections and asymptomatic infections in Figures 7 and 8.

**Figure 2:**
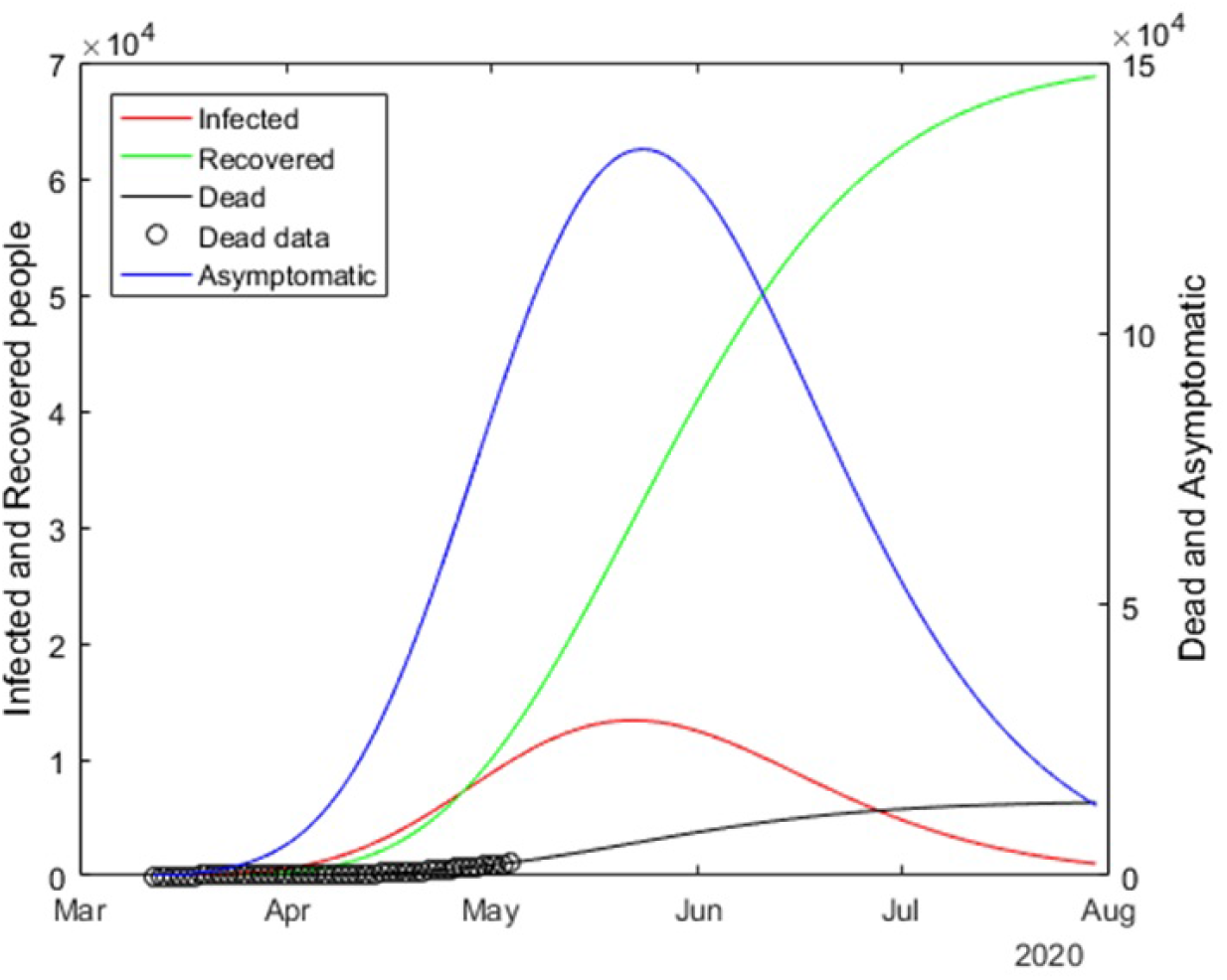
Graphs for the spread of COVID-19 in Mexico. The solid red line represents infected individuals, the solid blue line represents infected individuals but without any type of symptoms, finally, the solid black line represents the fatalities by the disease.

**Figure 3:**
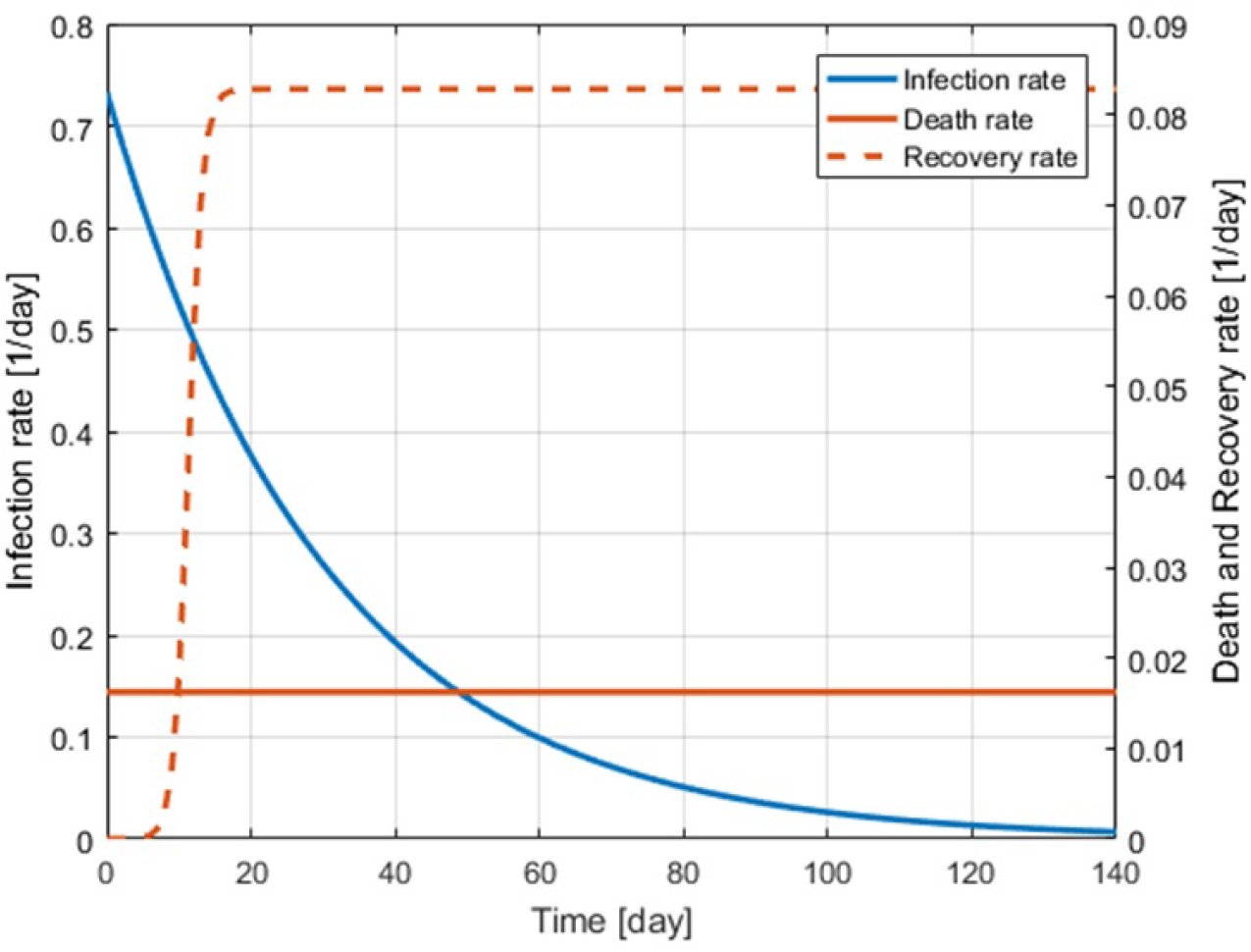
Best fit values of the infection, recovery and death rates as functions of time.

**Figure 4:**
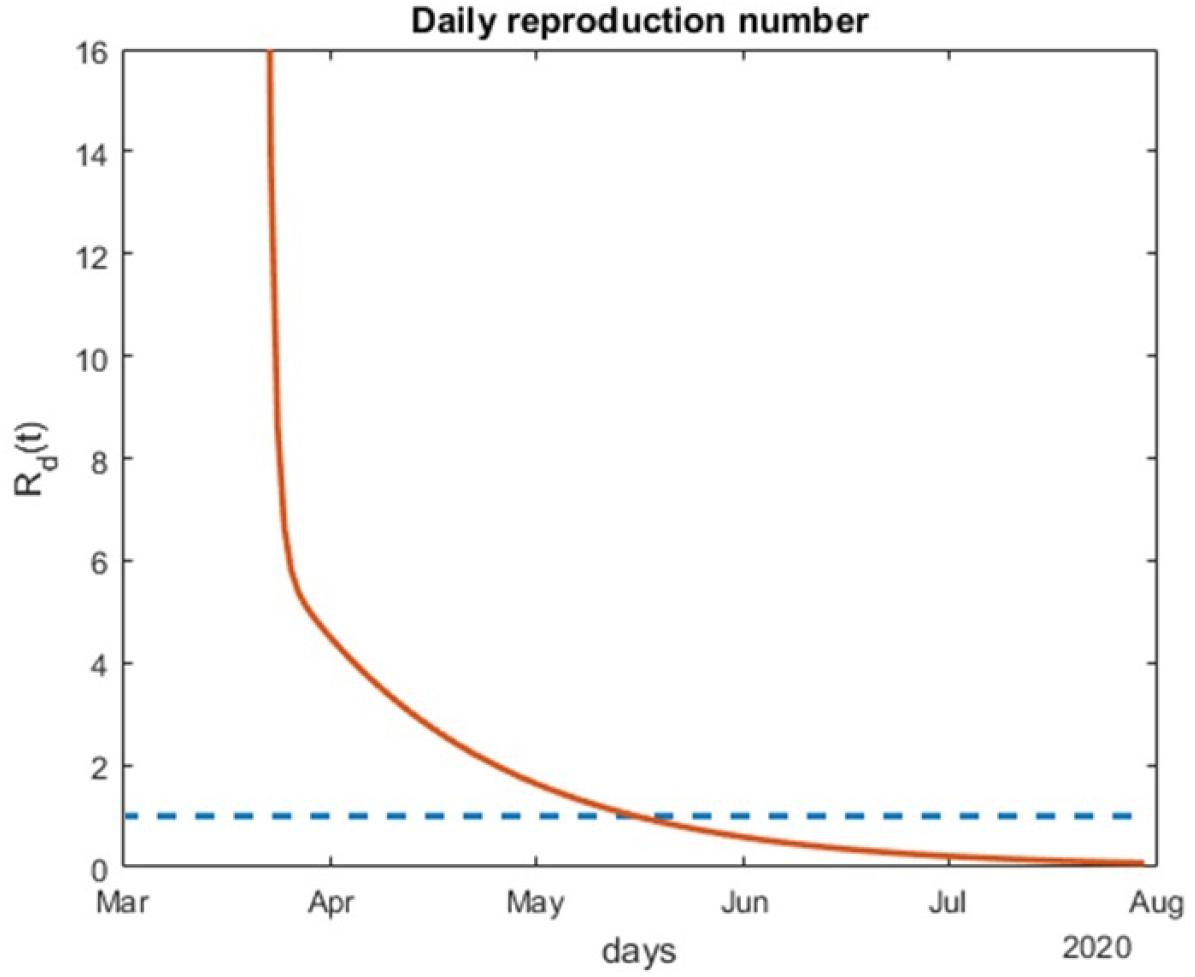
Variation of the effective daily reproduction number through time. The solid orange line represents the value of the reproduction number and the blue dotted line represents the value one.

**Figure 5:**
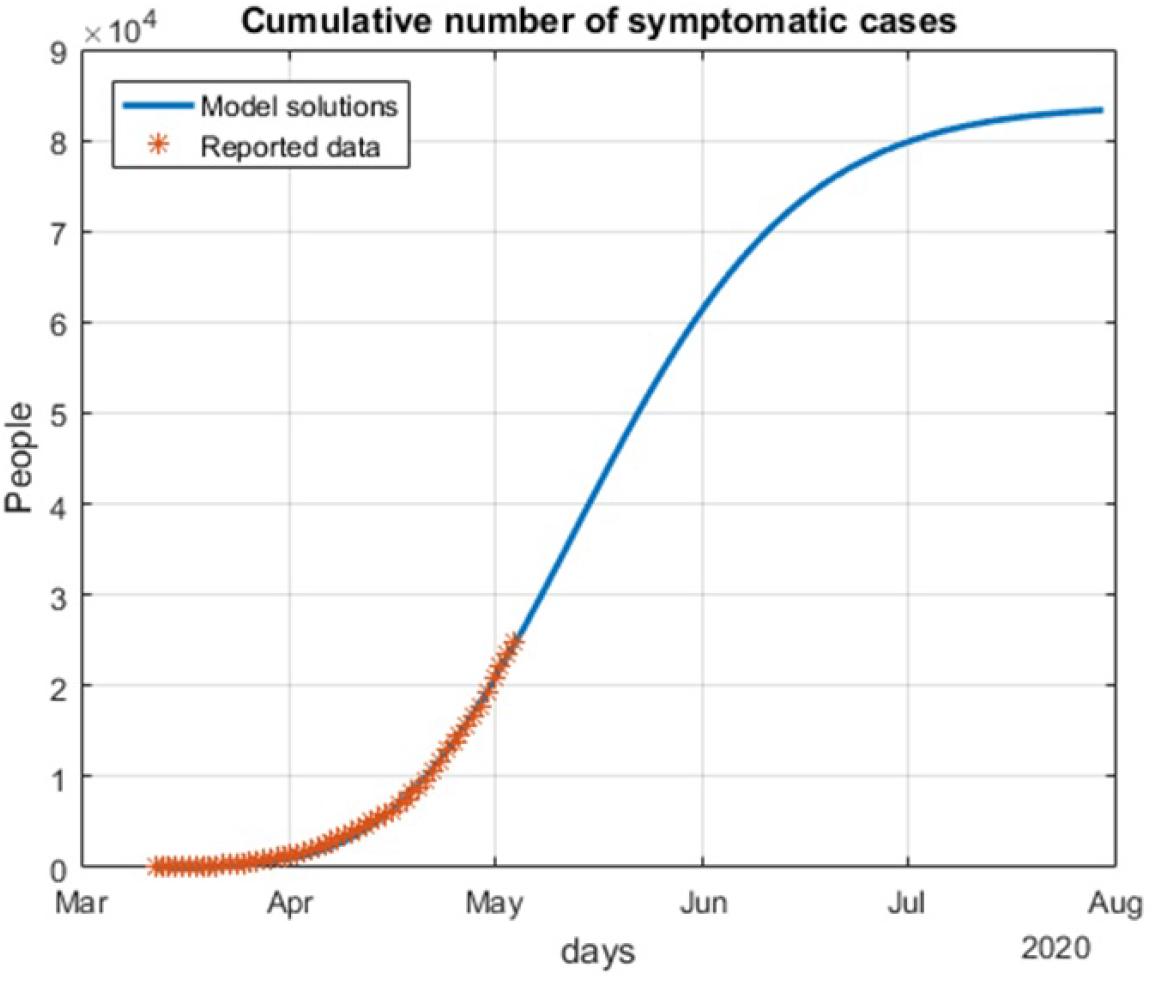
Cumulative number of symptomatic infected individuals (*I*(*t*) + *R_I_*(*t*) + *D*(*t*)) predicted by the model and reported cases.

**Figure 6:**
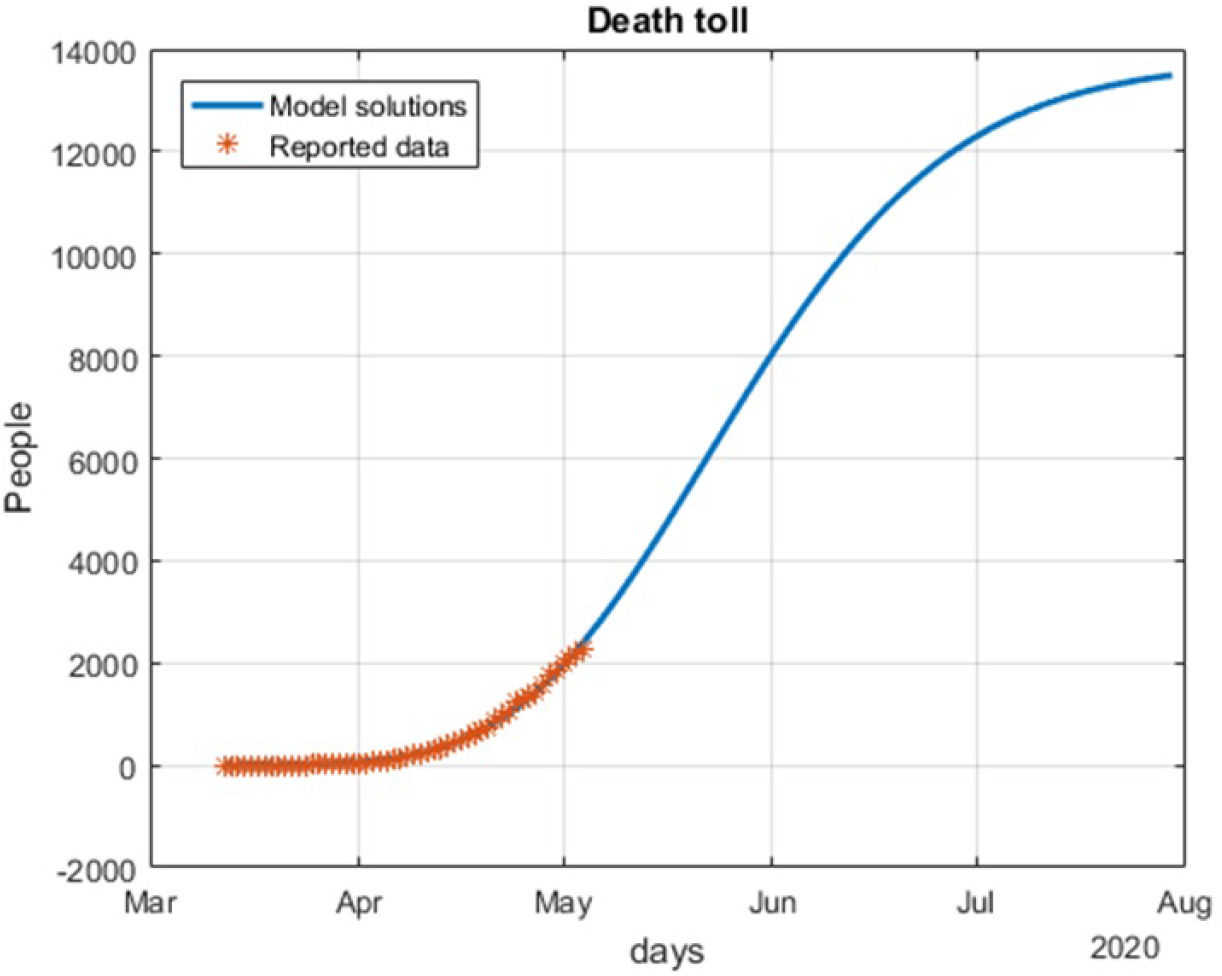
Death toll (*D*(*t*)) predicted by the model and reported number of fatalities.

**Figure 7:**
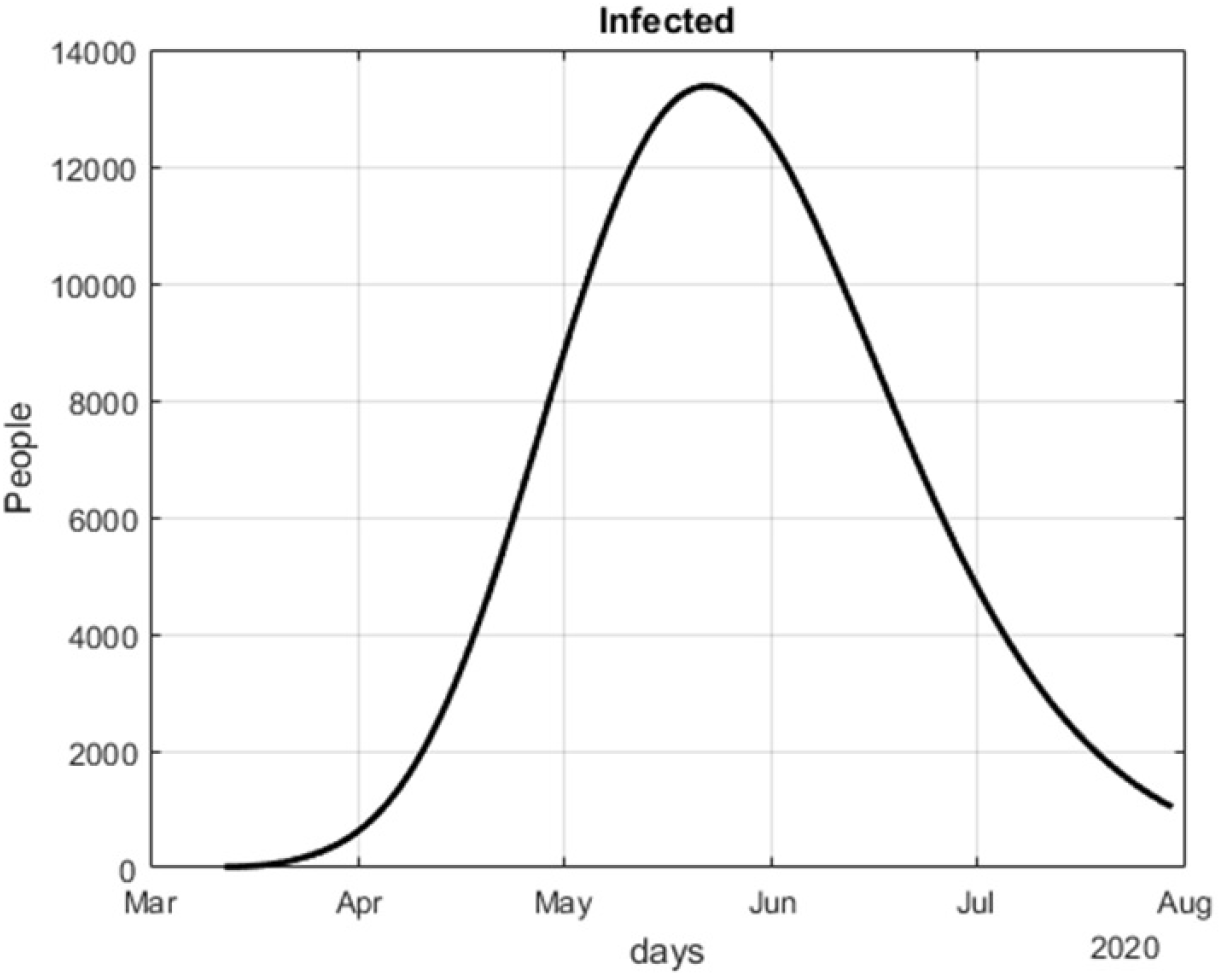
Number of infected cases (*I*(*t*)) predicted by the model.

**Figure 8:**
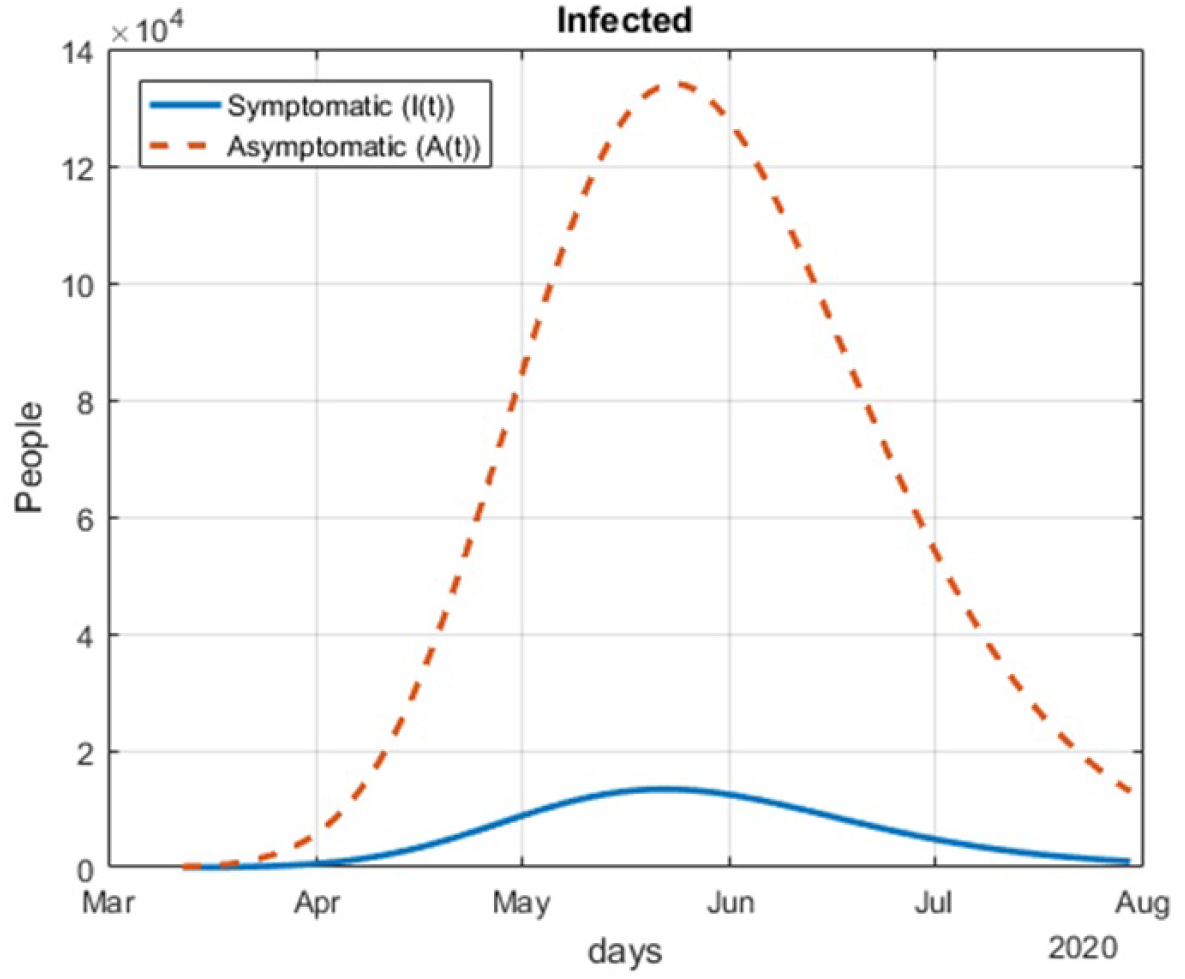
Number of infected cases (*I*(*t*)) and asymptomatic cases (*A*(*t*)) predicted by the model.

**Table 1:** Model parameters obtained from the best fit optimization for Mexico.

| Parameter | Best fit value for Mexico | Unit |
| --- | --- | --- |
| $\beta_0$ | 0.6052 | 1/day |
| $\beta_1$ | 0.01 | 1/day |
| $\tau_\beta$ | 30 | day |
| $\gamma_0$ | 0.0000 | 1/day |
| $\gamma_1$ | 0.0829 | 1/day |
| $\tau_\gamma$ | 11.2885 | day |
| $\delta_0$ | 0.0114 | 1/day |
| $\delta_1$ | 0.0088 | 1/day |
| $\tau_\delta$ | 76.1043 | day |
| $w$ | 0.2535 | 1/day |
| $p$ | 0.1 | — |

### 4.2 Heterogeneity of the outbreak of COVID-19 in two regions in Mexico

We evaluated the heterogeneity of the spread of COVID-19 in Mexico. We applied our SEIARD model for two important regions in Mexico: the Mexico Valley (Mexico City, State of Mexico and Morelos) and the Yucatan Peninsula (Campeche, Quintana Roo and Yucatan).

#### 4.2.1 Mexico Valley

First, we modeled the evolution of the outbreak of COVID-19 in the Valley of Mexico, which is shown in Figure 8. We adjusted the data provided by the Mexican Ministry of Health from March 12 to April 27 and simulated the daily cumulative cases for each state until August 2020. The outbreak in the Mexico Valley will be slightly different from the national perspective, and each state will behave differently.

Mexico City and State of Mexico (Figures 9A and 9B, respectively) have similar behavior, but in Morelos (Figure 9C) the peak of the infection moves to the right. The difference in the peak may be due to the separation of the capital of Morelos from Mexico City (86.6 km). Also, the connection between these states was cut off due to the confinement declared by the Ministry, which may be a reason for the difference between the outbreak of the infection in these states. Figure 10 shows the variation of the infection, recovery and death rates for the Mexico Valley. The rates between these states are different: Mexico City (the capital of Mexico, which has one of the two more important airports of the country) has a higher infection rate in the initial days of the simulation of our model by being close to 1. The death rate in Mexico City decreases considerably throughout the simulation, which means that this state may have less fatalities than other states in Mexico in the advanced stages of the outbreak. At the beginning, Morelos had a higher infection rate than Mexico City, but it decays later at roughly the same rate as Mexico City. At first the recovery rate in Morelos was very low and the death rate was high, this means that Morelos was impacted at first with severe cases.

**Figure 9:**
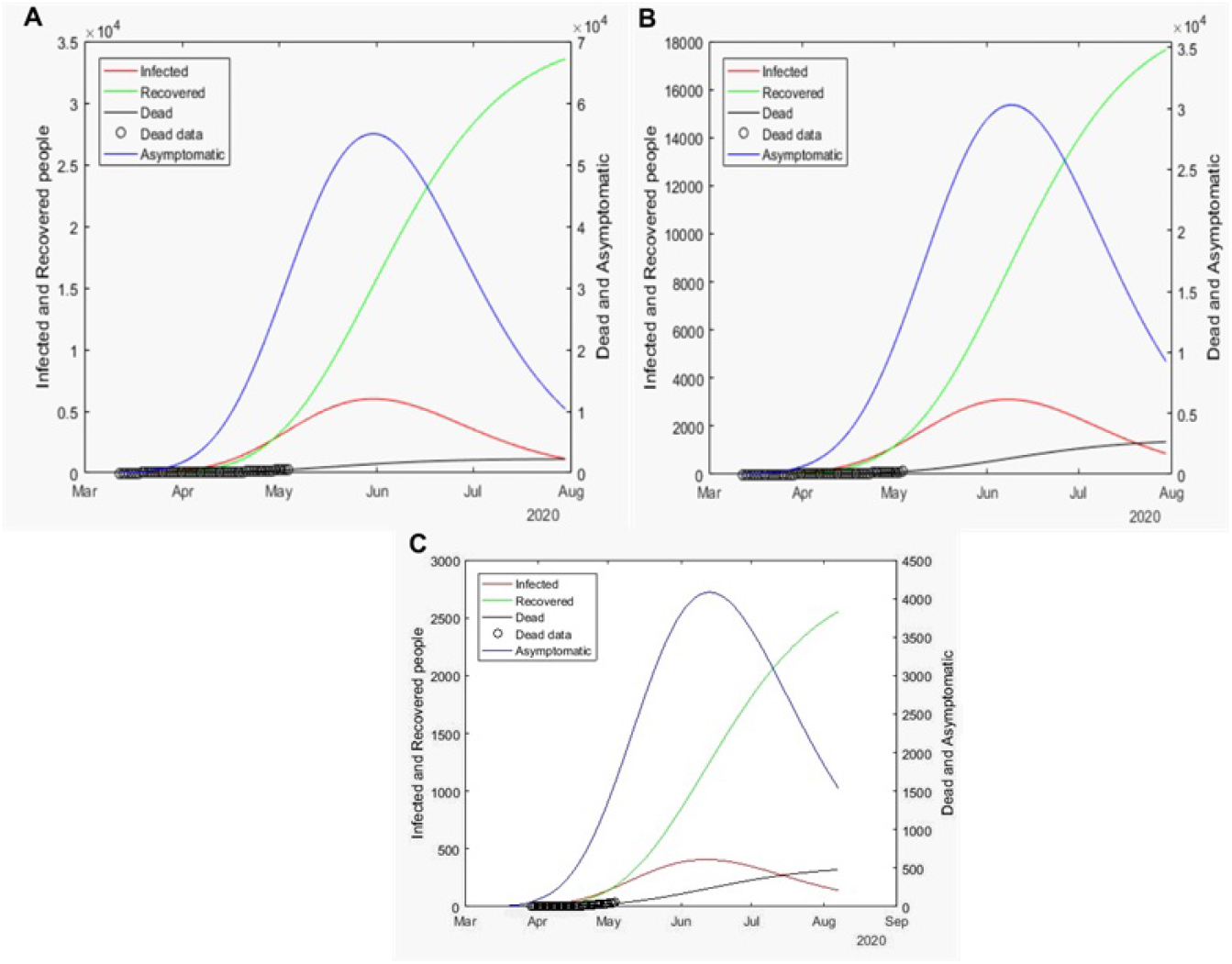
Graphs for the spread of COVID-19 in the Valley of Mexico. Solid red lines represent infected individuals, solid blue lines represent infected individuals but without any type of symptoms, finally, solid black lines represent the fatalities by the disease. A represents the outbreak for Mexico City, B represents the outbreak for the state of Mexico and C for Morelos.

**Figure 10:**
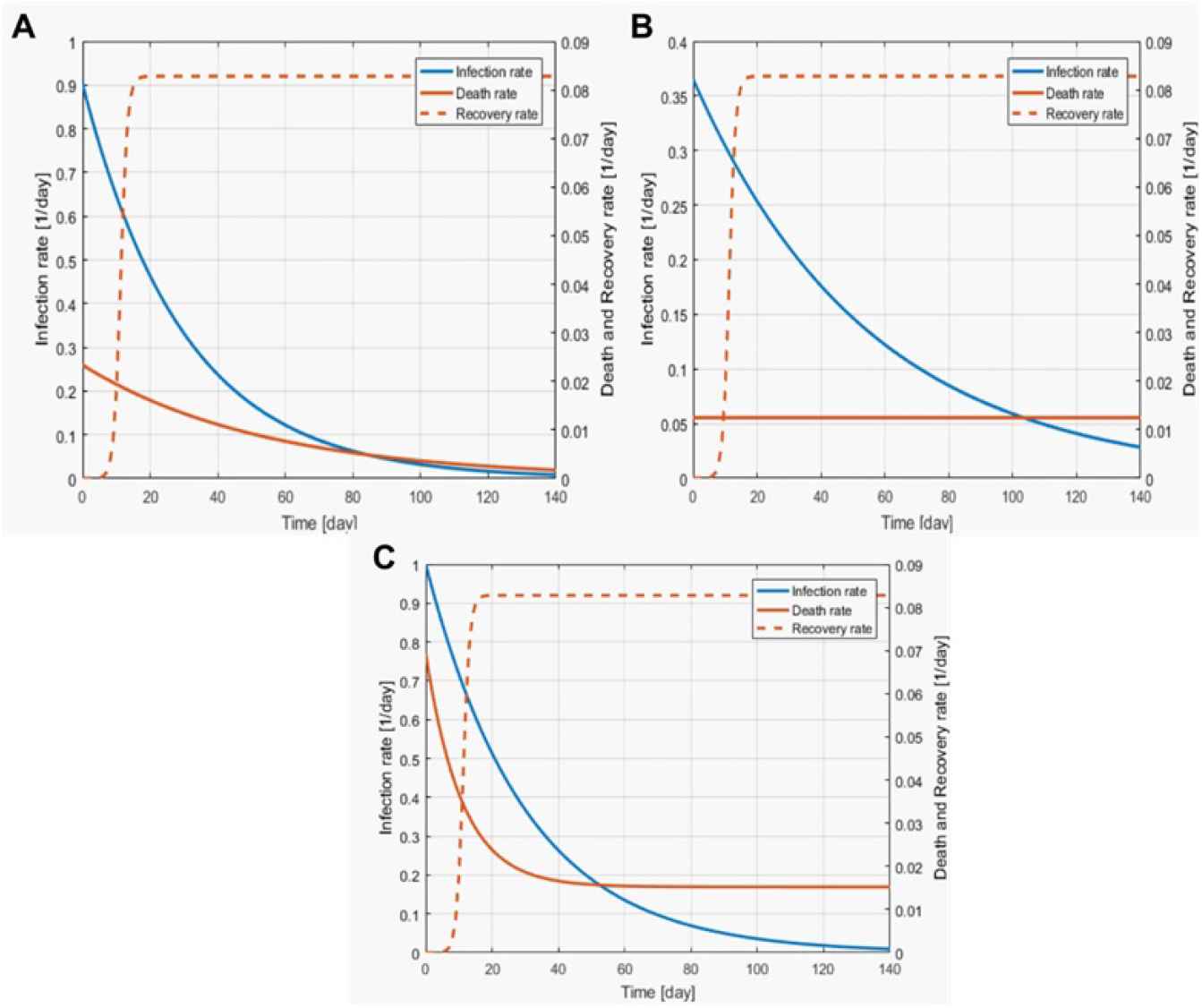
Best fit parameters of the infection, death and recovery rates as functions of time for the Valley of Mexico. Solid orange line is the death rate, dotted orange line the recovery rate and the solid blue line the infection rate. A depicts the rate for Mexico City, B for the State of Mexico and C for Morelos.

Once times passes, the recovery rate increases due to more milder cases and the death rate decreases as well. The daily reproduction number (Figure 11) in Mexico City decreases at roughly the same rate as in Morelos and State of Mexico. We believe that Mexico City should be treated differently to the other states in the Mexico Valley since this city has a much higher influence of transients daily. This is due to citizens from the surrounding areas coming to this city to work, creating a larger population that comprises not only native citizens, but citizens from State of Mexico and Morelos as well. In Figures 12-15, we carried out the simulation with the best fit parameters. We show a comparison of the cumulative number of infections: as we can see, Mexico City (Figure 12A) will have the greatest number of infections within the pandemic of COVID-19, the state of Mexico (Figure 12B) will be the second state with most infections.

**Figure 11:**
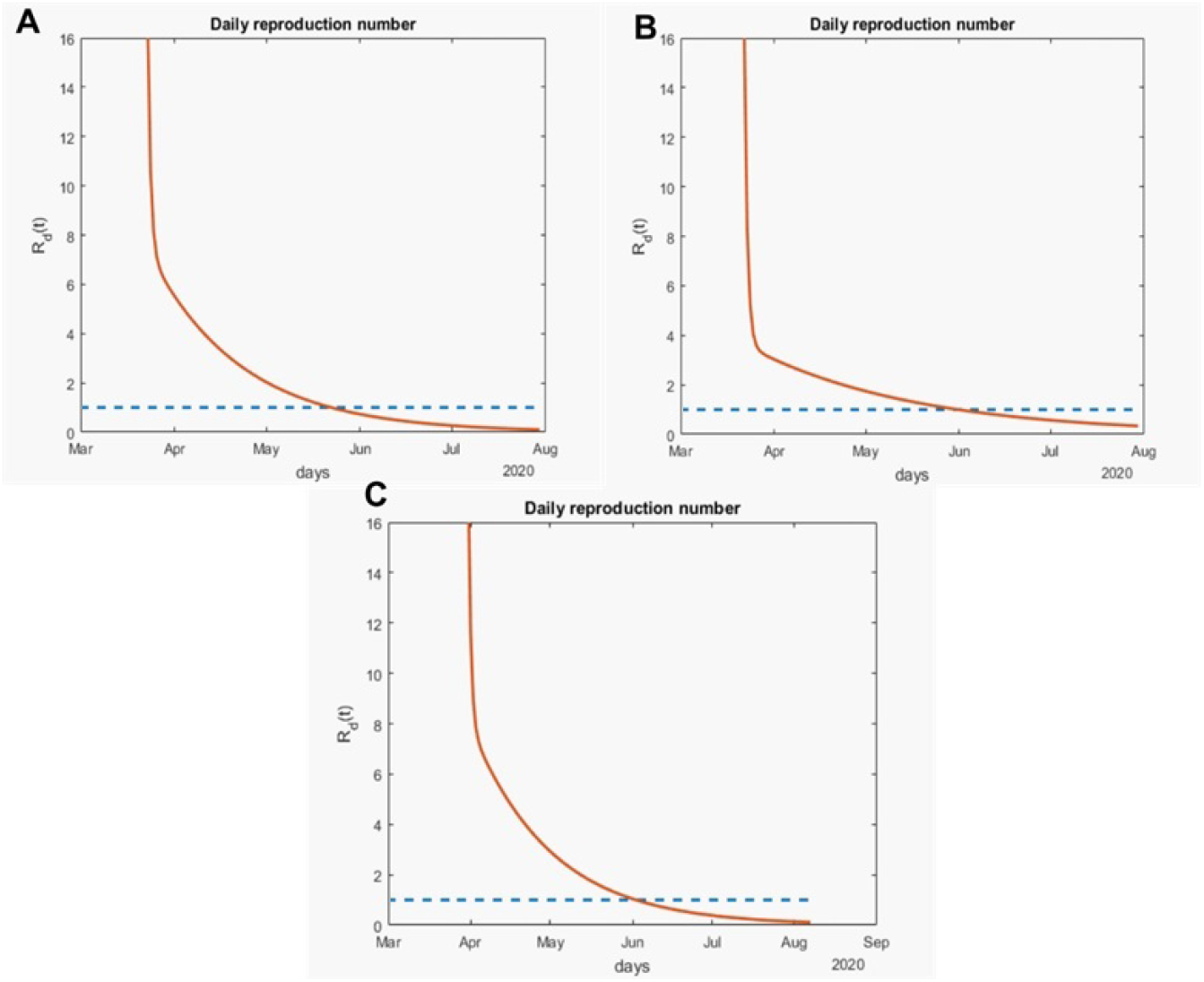
Variation of the effective daily reproduction number through time for the Mexico Valley. The solid orange line represents the value of the reproduction number and the blue dotted line represents the value one. A represents the value for Mexico City, B for the State of Mexico and C for Morelos.

**Figure 12:**
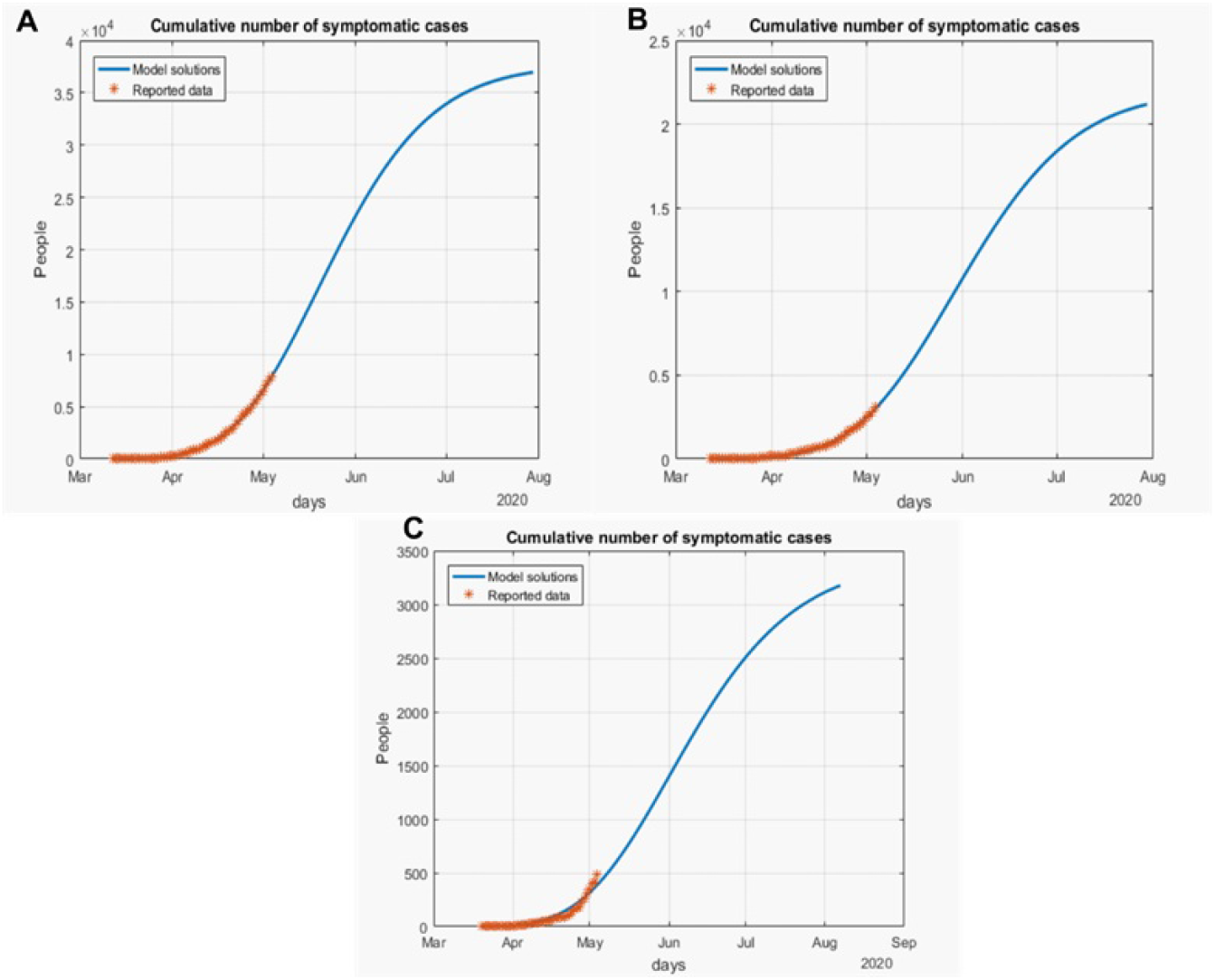
Cumulative number of symptomatic infected individuals *C*(*t*) = *I*(*t*) + *R_I_*(*t*) + *D*(*t*) predicted by the model and reported cases for the Mexico Valley. A represents Mexico City, B the State of Mexico and C Morelos.

**Figure 13:**
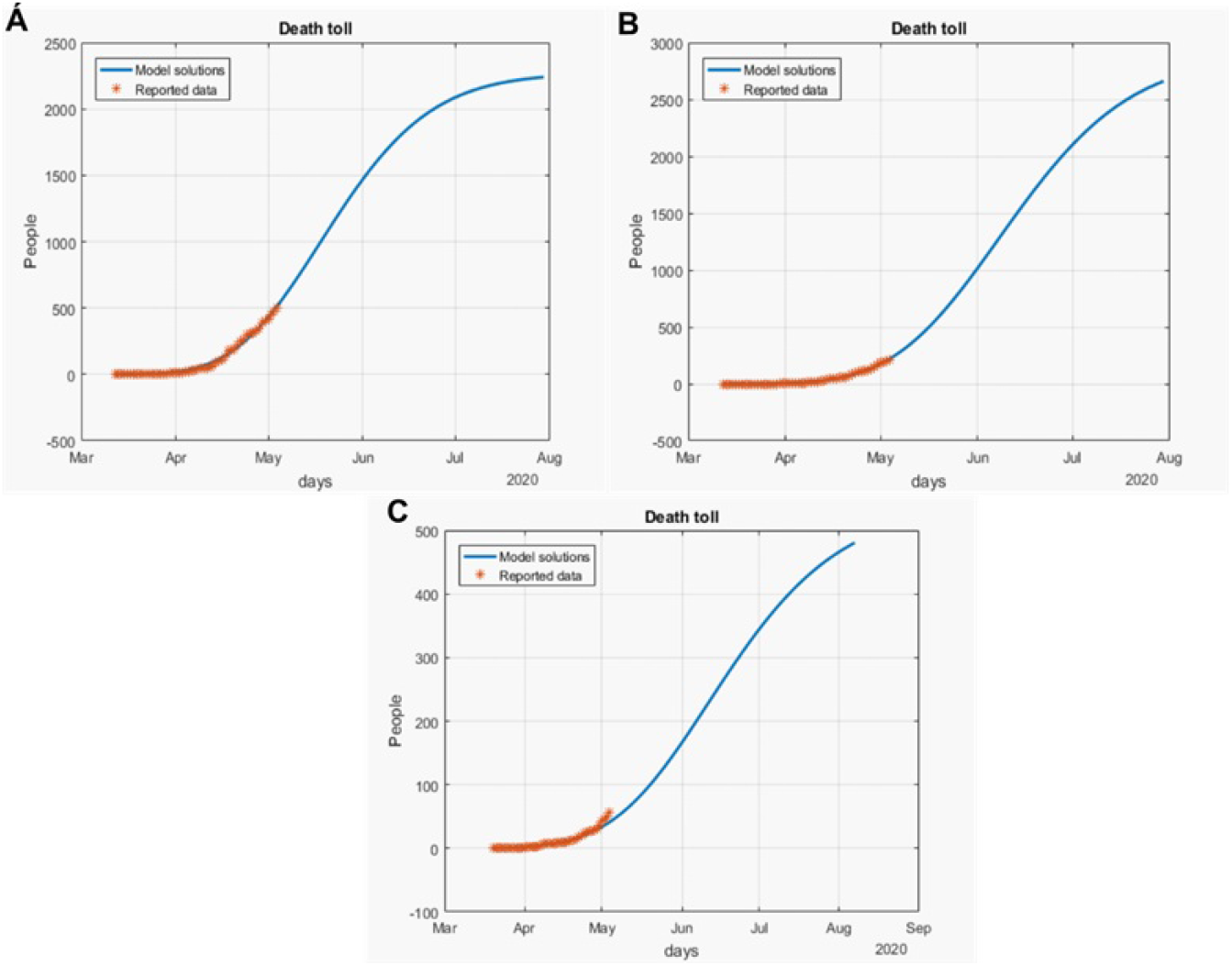
Death toll (*D*(*t*)) predicted by the model and the reported number of fatalities for the Mexico Valley. A represents the fatalities for Mexico City, B for the State of Mexico and C for Morelos.

**Figure 14:**
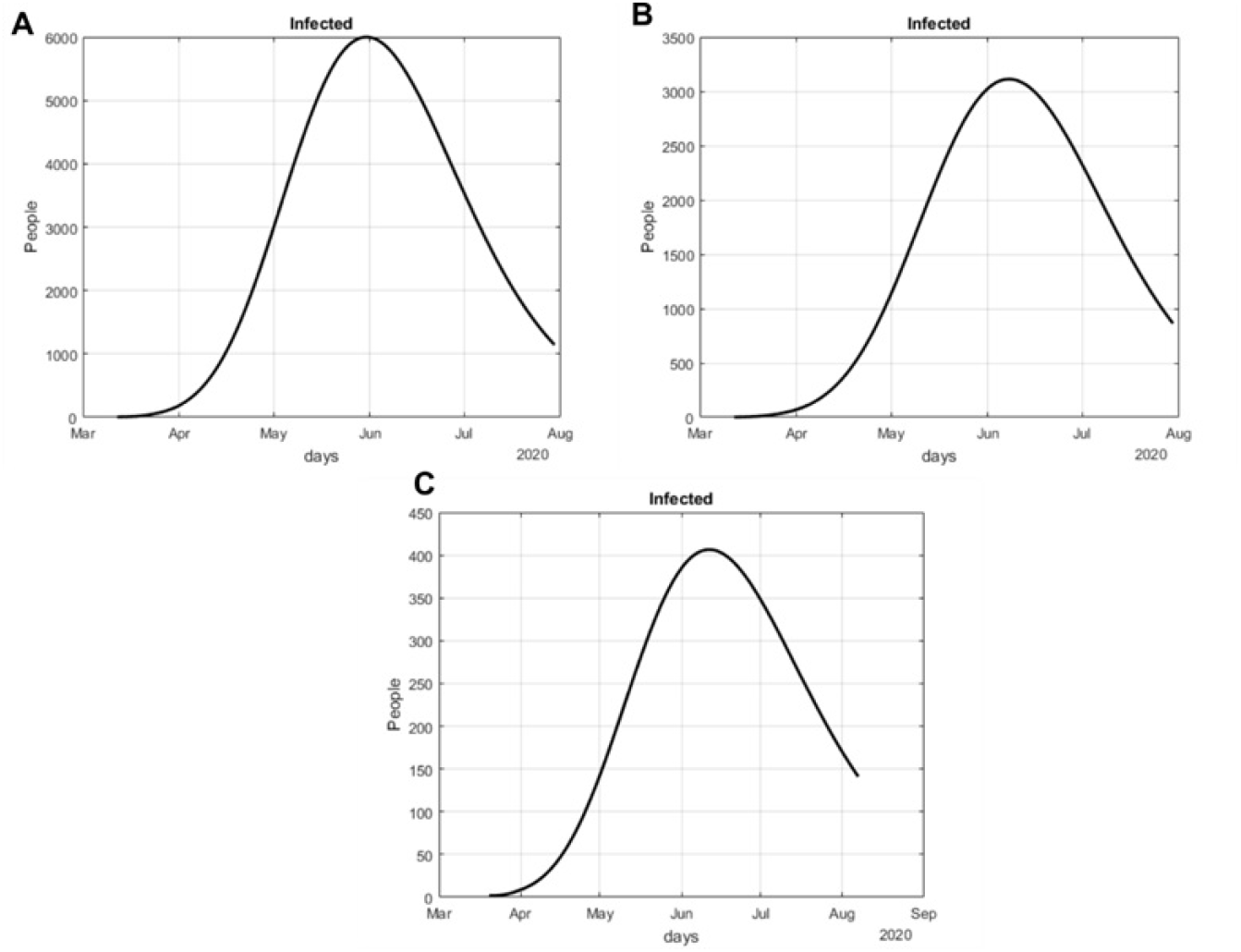
Number of infected cases (*I*(*t*)) predicted by the model of the Mexico Valley. A represents Mexico City, B State of Mexico and C Morelos.

**Figure 15:**
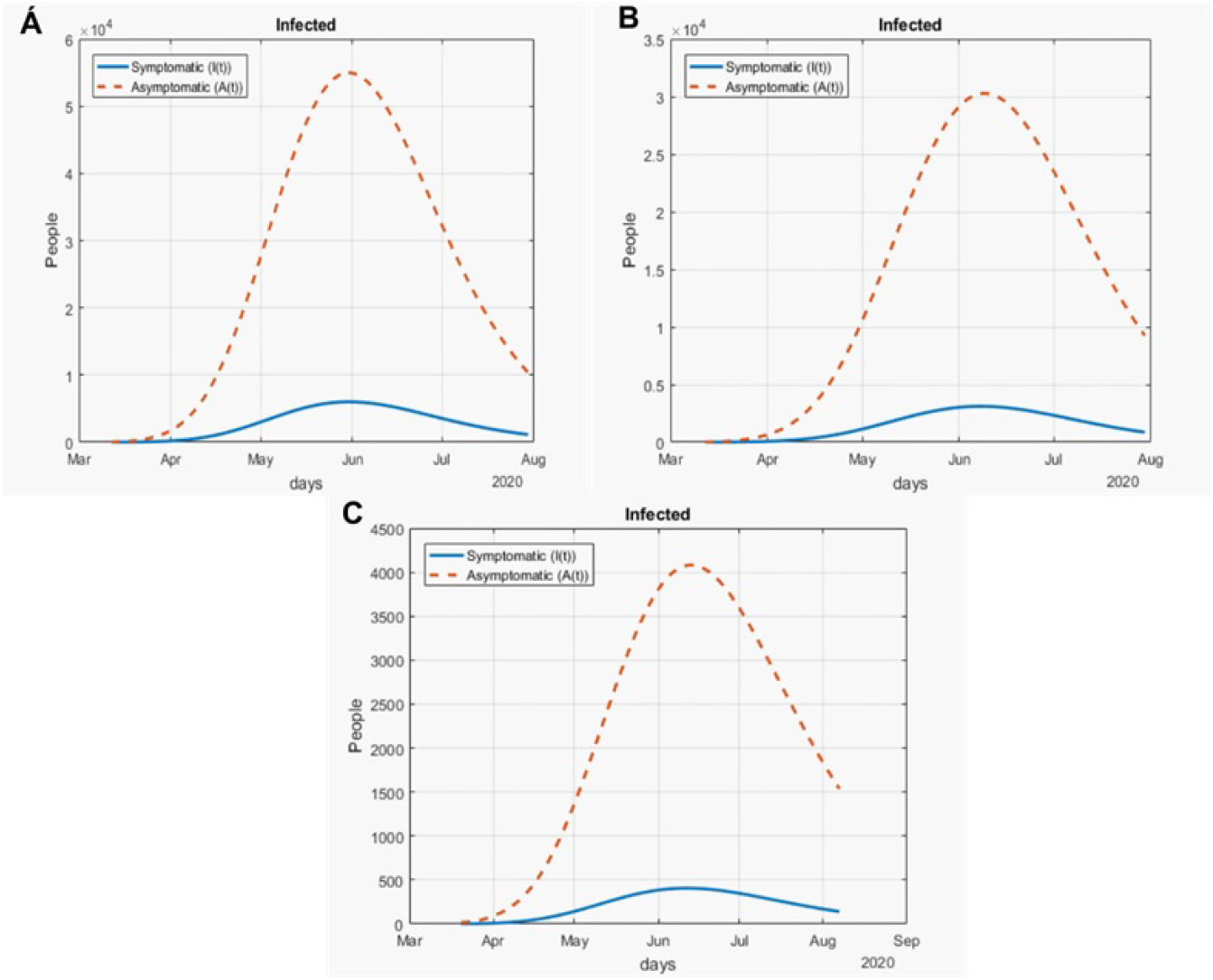
Number of infected cases (*I*(*t*)) and asymptomatic cases (*A*(*t*)) predicted by our model for the Mexico Valley. A represents Mexico City, B State of Mexico and C Morelos.

Mexico City will have most of the fatalities caused by the virus, followed by the State of Mexico, which has the cities with higher population than the rest of the states (Figure 13) with the reported data. We also plot the number of active symptomatic infections and asymptomatic infections in Figures 14 and 15. Asymptomatic individuals are of great importance as they present a much higher number with respect to symptomatic individuals, by activating the “Jornada de Distanciamiento Social”, the number of symptomatic infections has been reduced and the curve of this individuals has been flattened as well.

#### 4.2.2 Yucatan Peninsula

Now, we evaluate the outbreak in the Yucatan Peninsula. We reviewed Campeche, Yucatan and Quintana Roo; the outbreaks are depicted in Figure 16. The peak of the infection in Campeche (Figure 16A) and Quintana Roo (Figure 16B) will be around the last week of May. For the state of Yucatan (Figure 16C), it will be during the month of June, the outbreak of COVID-19 is still starting as we will see in the next figures.

**Figure 16:**
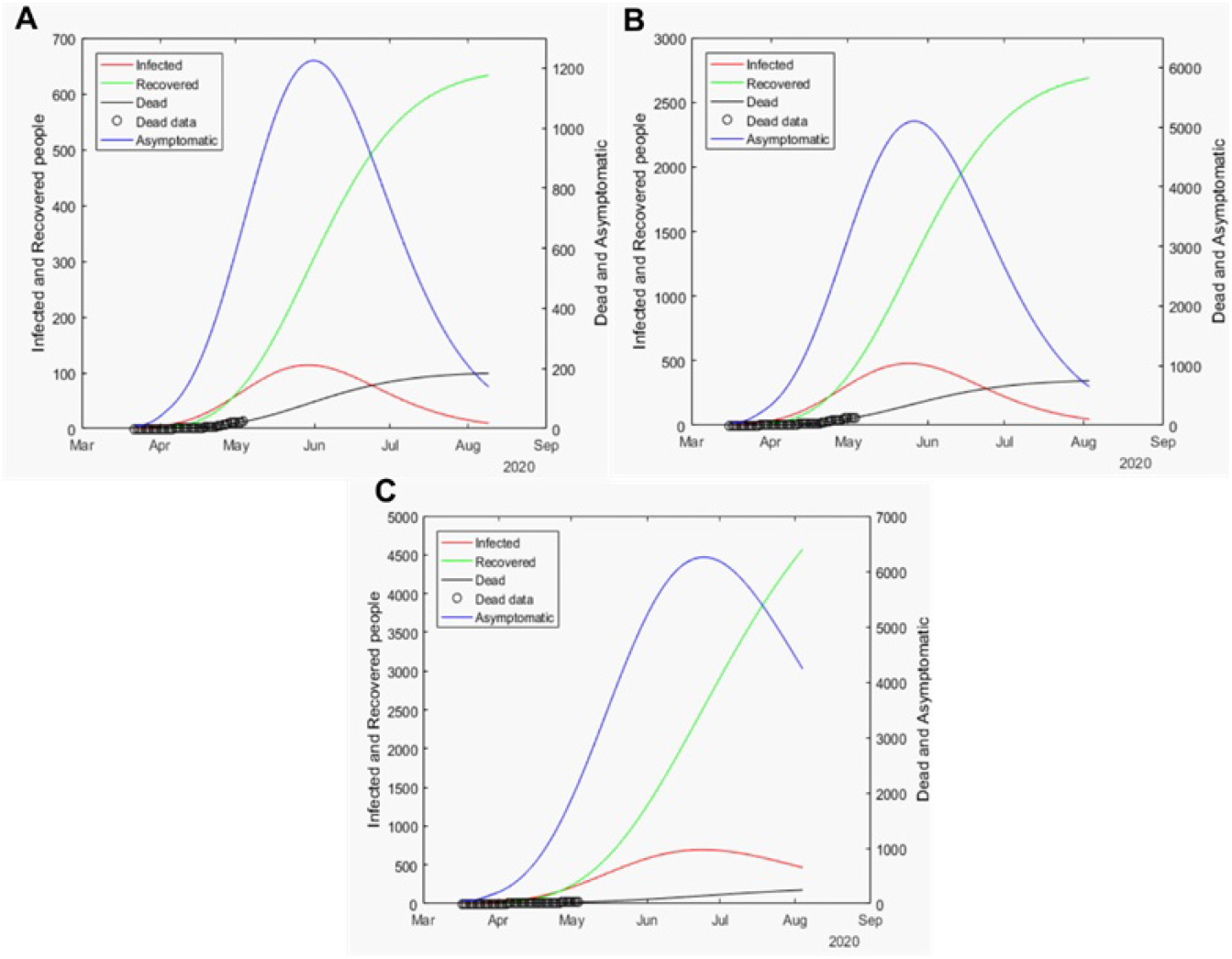
Graphs for the spread of COVID-19 in the Yucatan Peninsula. Solid red lines represent infected individuals, solid blue lines represent infected individuals but without any type of symptoms, finally, solid black lines represent the fatalities by the disease. A represents the outbreak for Campeche, B represents the outbreak for Quintana Roo and C for Yucatan.

Yucatan has the highest infection rate of the Peninsula with a value equal to one at the beginning of the simulation (Figure 17C). Campeche (Figure 17A) starts with an infection rate of 0.45, while Quintana Roo (Figure 17B) starts with an infection rate of 0.5; the decay occurs at similar rates for these two states. The recovery rate for the Yucatan Peninsula behaves the same between the states. In the first days of the infection, there will be more severe cases and then a mixture of severe and mild cases. By day twenty, the recovery rate for the three states will remain constant. At the beginning of the simulation, Yucatan (Figure 17C) has a higher death rate compared to the other states, but later it decays rapidly to less than 0.01. The death rates for Campeche (Figure 17A) and Quintana Roo (Figure 17B) remain constant through most of the simulation with values slightly higher than 0.02.

**Figure 17:**
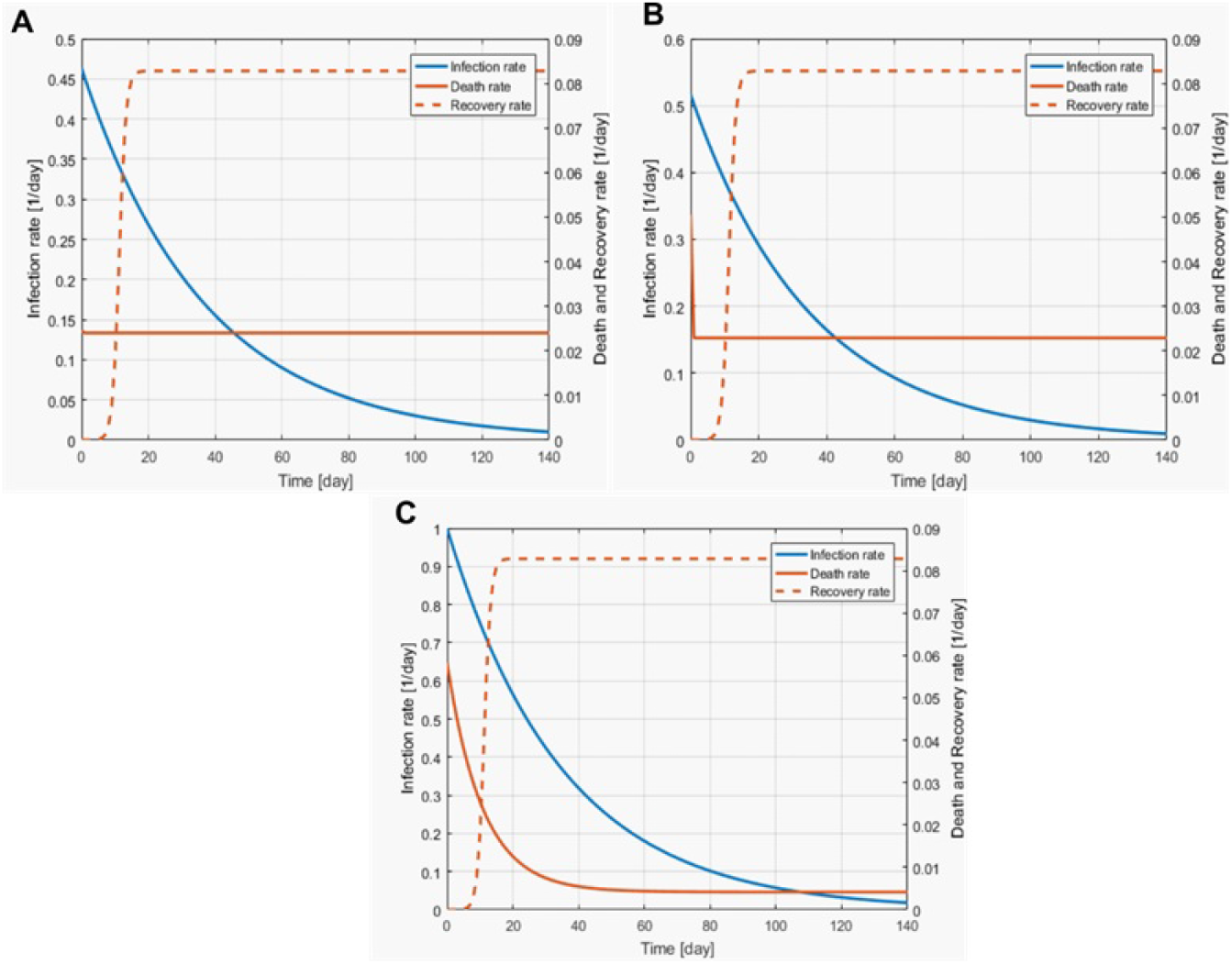
Best fit parameters of the infection, death and recovery rates as functions of time for the Yucatan Peninsula. Solid orange line is the death rate, dotted orange line the recovery rate and the solid blue line the infection rate. A depicts the rate for Campeche, B for Quintana Roo and C for Yucatan.

Regarding the daily reproduction number, Campeche and Quintana Roo (Figures 18A and 18B, respectively) will have the same behavior: by early April, both will have a value for *R_d_*(*t*) of 4 and they will enter phase two with a value smaller than 3. During the month of May, the reproduction number will become smaller than one. Yucatan has the same behavior, but the reproduction number will be smaller than one by early June, this state should evaluate how well they are staying at their houses to try to diminish *R_d_*(*t*) at the same velocity than the rest of the states of the Yucatan Peninsula. We thought Quintana Roo (Figure 19B) was going to have a higher number of infected individuals, but Yucatan will have much more than the other states (Figure 19C). Campeche will have slightly more than 800 infected individuals. The death toll will be higher in Quintana Roo (Figure 20B) and then Yucatan (Figure 20C). Alarmingly, in Campeche roughly 20 percent of the infected will die by complications product of COVID-19. This behavior is of great importance, but we cannot know in detail why this is happening in this state. Finally, we can see in Figure 22 the simulation of the symptomatic and asymptomatic cases, which shows that the Yucatan peninsula is indeed flattening the curve.

**Figure 18:**
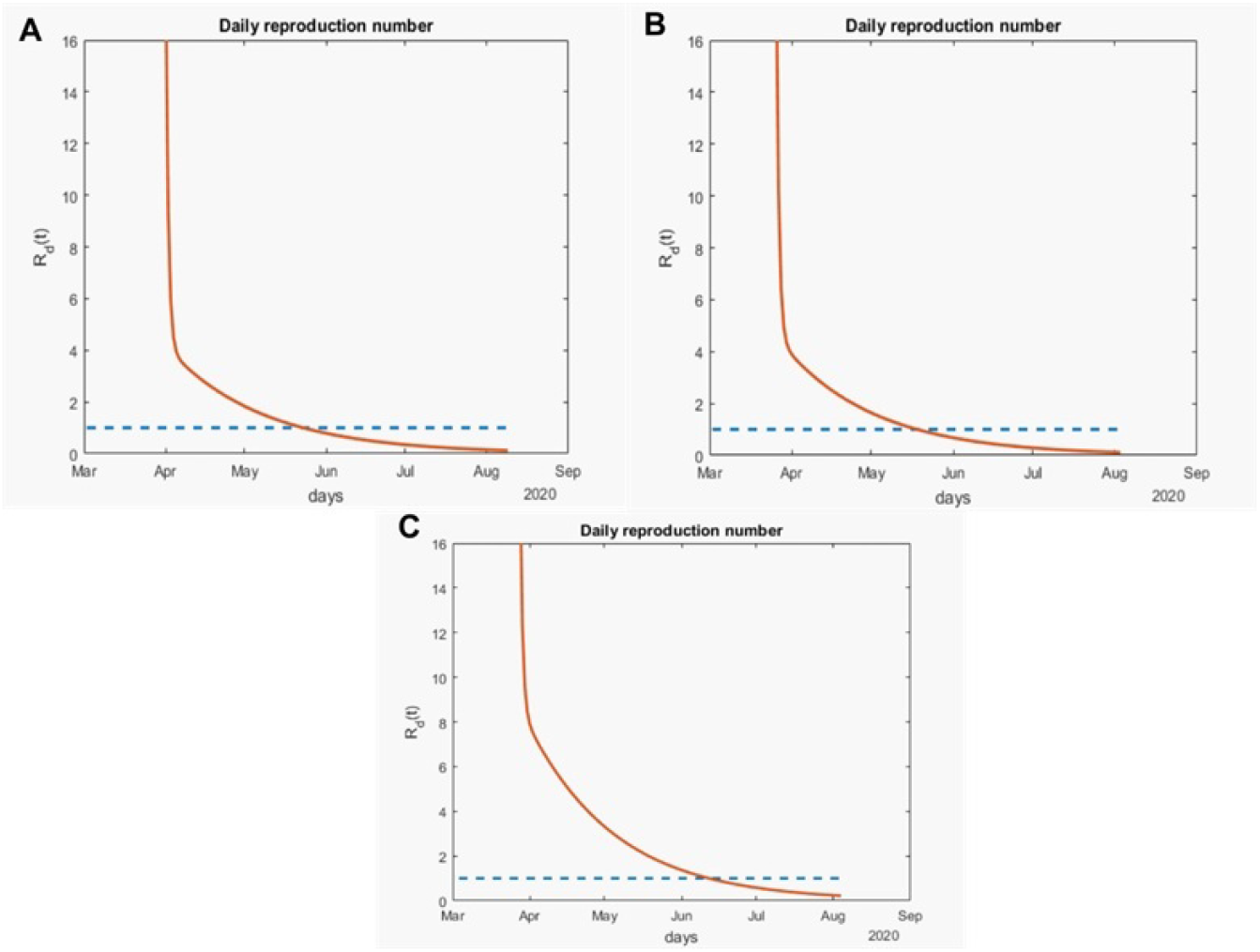
Variation of the effective daily reproduction number throughout time for the Yucatan Peninsula. The solid orange line represents the value of the reproduction number and the blue dotted line represents the value one. A represents the value for Campeche, B for Quintana Roo and C for Yucatan.

**Figure 19:**
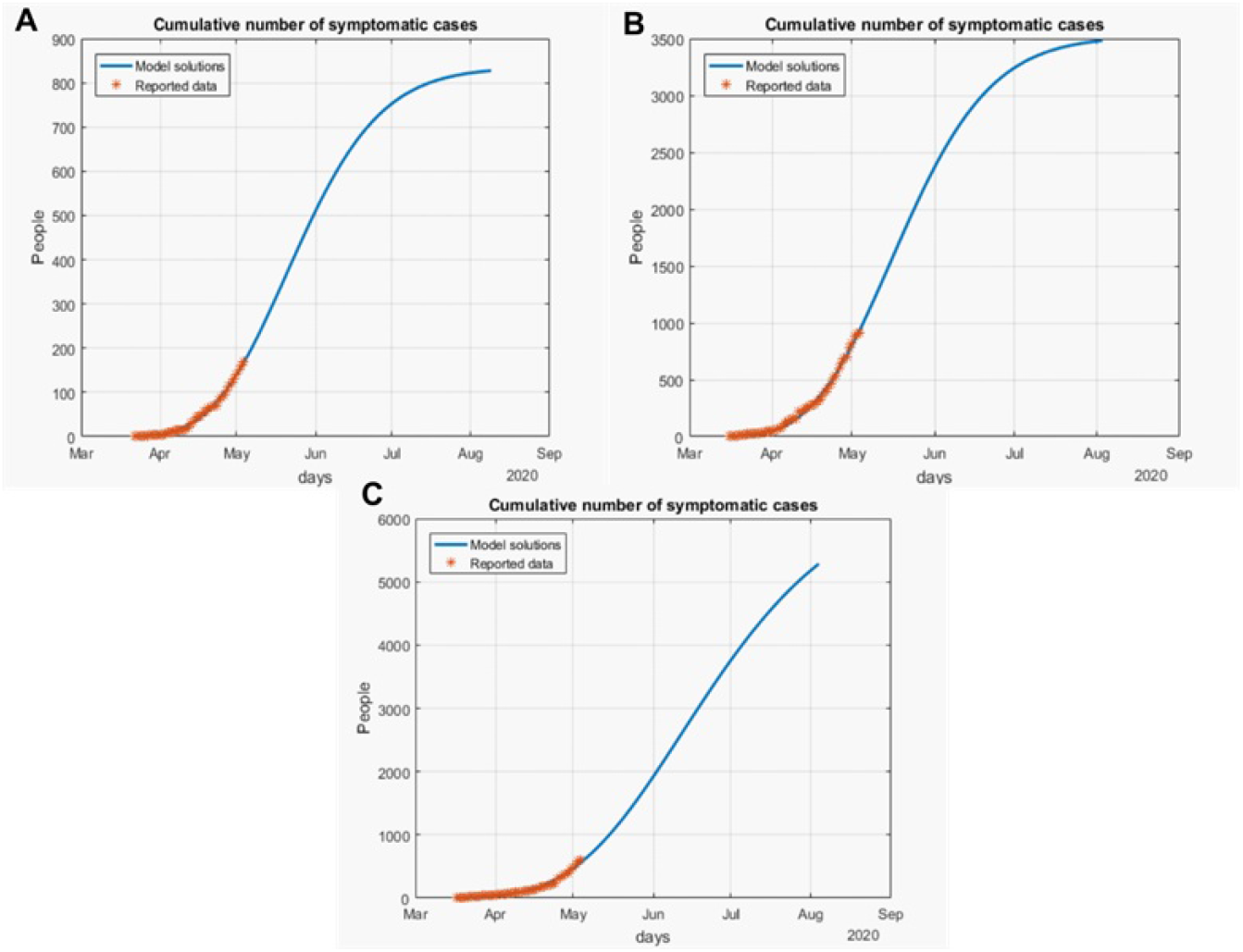
Cumulative number of symptomatic infected individuals *C*(*t*) = *I*(*t*) + *R_I_*(*t*) + *D*(*t*) predicted by the model and reported cases for the Yucatan Peninsula. A represents Campeche, B Quintana Roo and C Yucatan.

**Figure 20:**
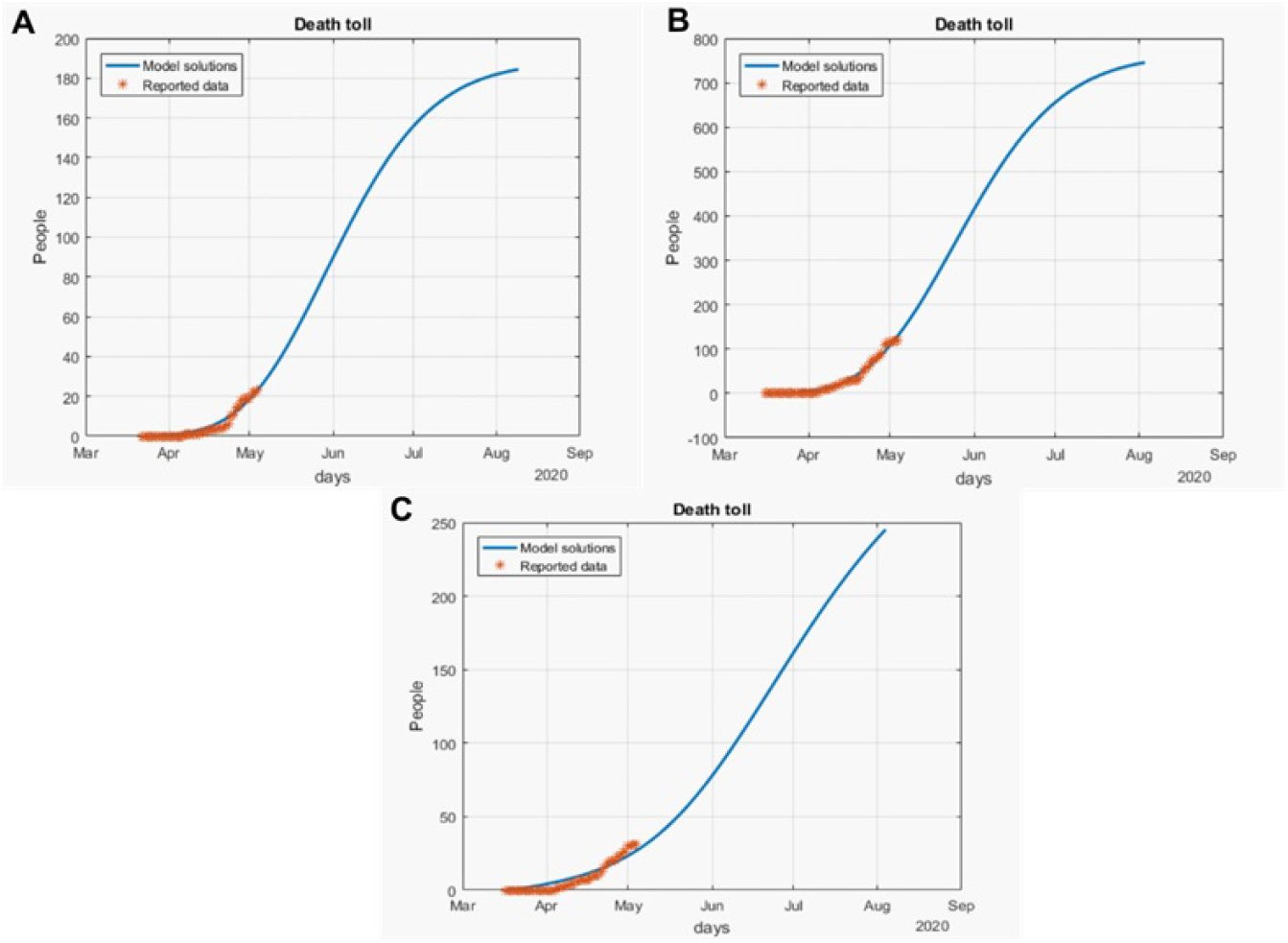
Death toll (*D*(*t*)) predicted by the model and the reported number of fatalities for the Yucatan Peninsula. A represents the fatalities for Campeche, B for Quintana Roo and C for Yucatan.

**Figure 21:**
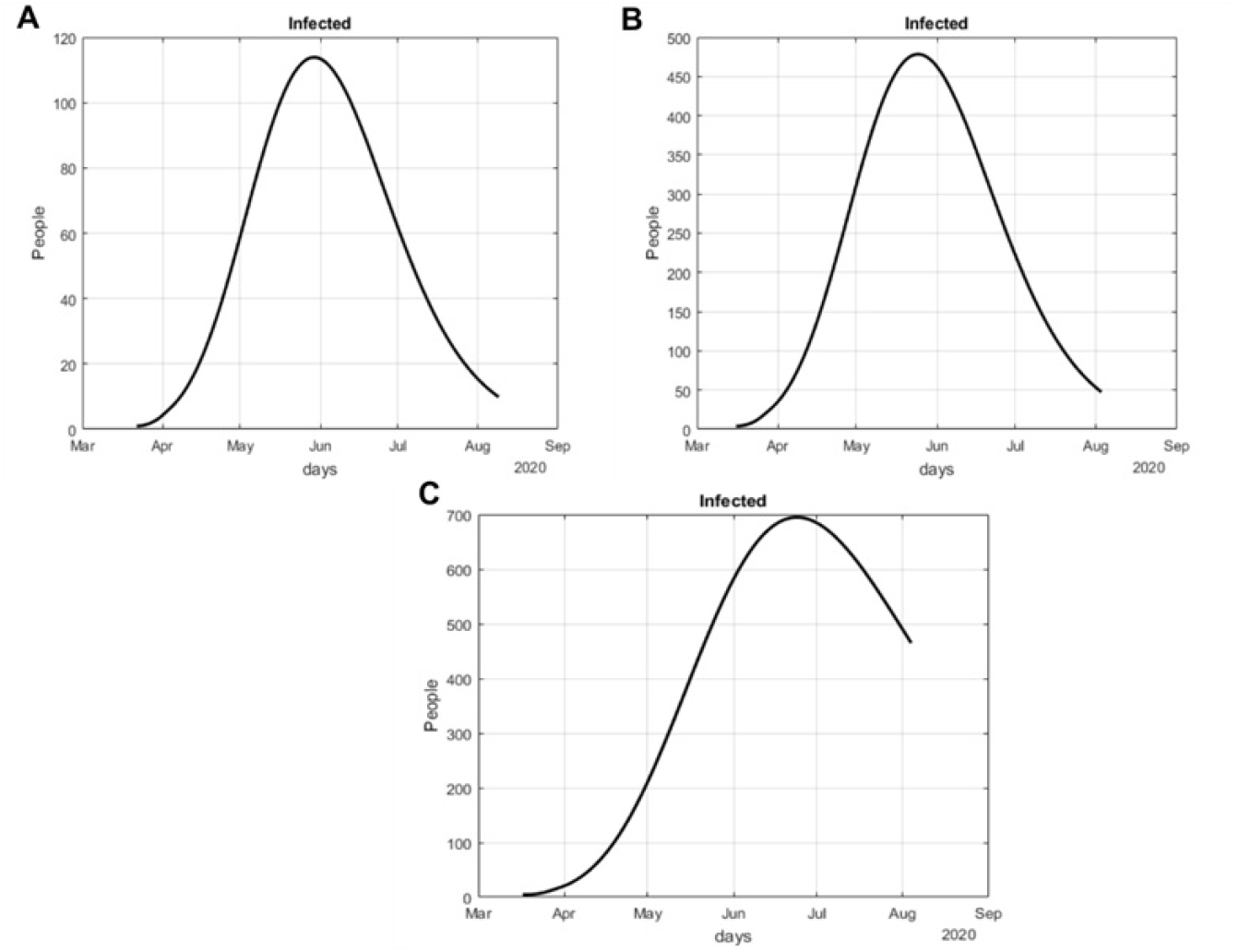
Number of infected cases (*I*(*t*)) predicted by the model of the Yucatan Peninsula. A represents Campeche, B Quintana Roo and C Yucatan.

**Figure 22:**
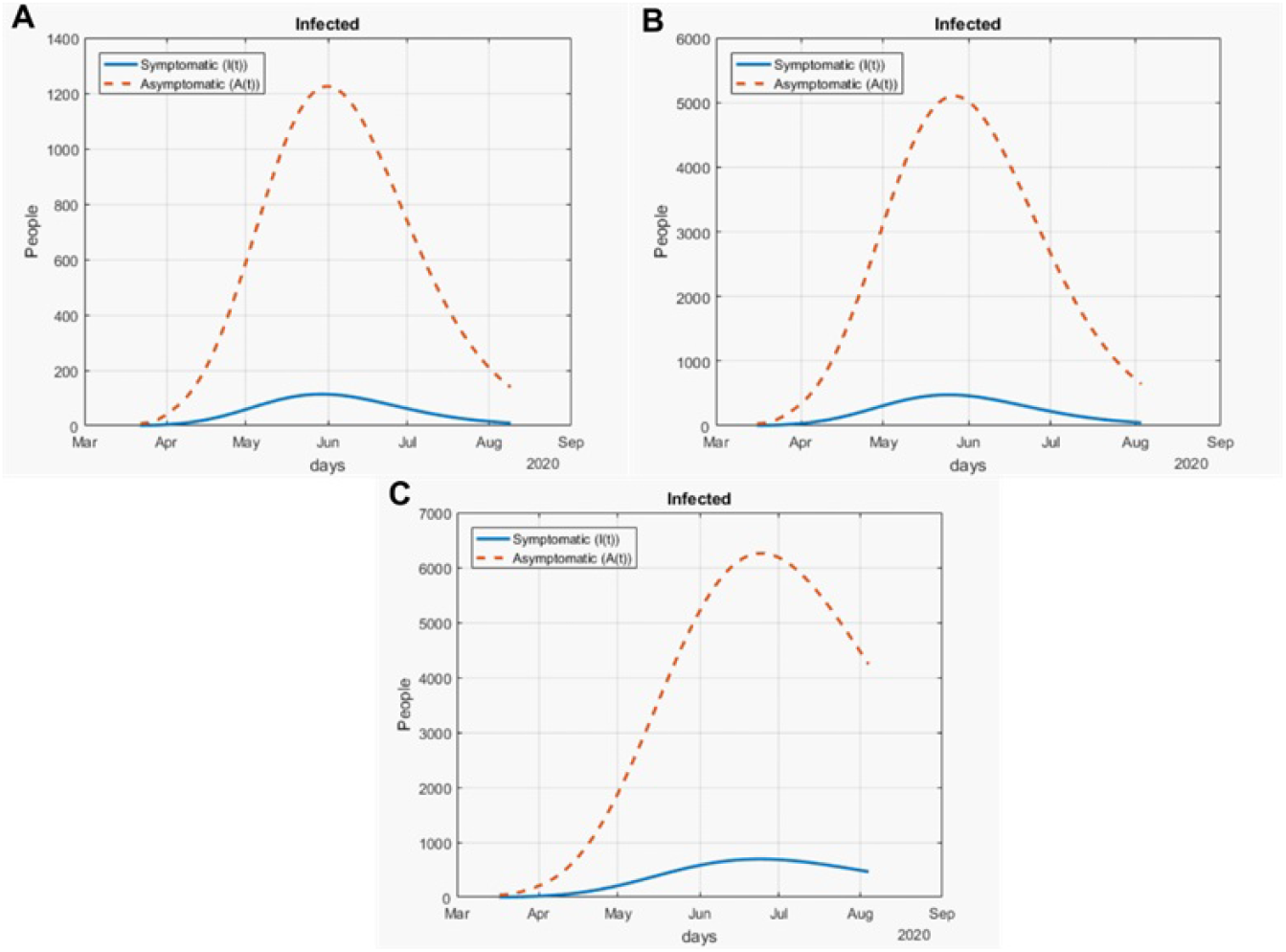
Number of infected cases (*I*(*t*)) and asymptomatic cases (*A*(*t*)) predicted by our model for the Yucatan Peninsula. A represents Campeche, B Quintana Roo and C Yucatan.

## 5 Discussion

During the outbreak of a pandemic where the transmission is from infected individuals to healthy individuals, the use of mathematical models as a forecast is of great importance. By this approach, authorities can plan a health care program and control the spread even with limited resources. In this work, we formulated and analyzed a compartmental mathematic epidemic model to simulate the outbreak of the virus in mainland Mexico and important regions of said country.

Our mathematical model was a data driven analysis, using data publicly available from our Ministry of Health, which are updated daily. Our model incorporates two compartments that are of the utmost importance: exposed and asymptomatic. By incorporating the exposed subpopulation, we are modeling the latency period, and the asymptomatic subpopulation incorporates the individuals that do not present any type of symptoms but have the ability to spread the virus and infect more individuals. Both tools are important to understand the size and time of the outbreak.

The results of our numerical solutions would let us compare how different the outbreak will be in the two studied regions. For overall Mexico, we used the cumulative infections for all 32 states. The Valley of Mexico will be the region with most infected individuals and deaths, because this region concentrates 22% of the Mexican population. Nevertheless, there is still heterogeneity in the Valley of Mexico: Mexico City and the State of Mexico present a similar behavior, but in Mexico City they will be more infected individuals with or without symptoms. Even though control measures like social distancing were applied and the curve of infected individuals is flattened, there will be a high number of infected individuals with symptoms. This being said, Mexico should focus their effort in applying hospital interconnection and have the sufficient resources to help all individuals that will develop more severe respiratory symptoms.

The Yucatan Peninsula has the same behavior on how the number of infections is extended throughout time (the curve is flattened), but the number of infected individuals is different. We recommend that Mexico should focus their actions in supplying medical stock to Quintana Roo, because this region will have more infected individuals, as well as Yucatan. Campeche, on the other hand, has an alarming behavior, this state will have less infected individuals from the Yucatan Peninsula, but they will have a very high death rate compared with their number of infected individuals. Mexico should be concerned about why in this state the number of fatalities is high compared to their size of population.

We believe Mexican authorities applied the control measures like social distancing and quarantine at the right time, and the number of infections in the regions is the expected based on their population size. It is very important to mention that even though Mexico succeeded in flattening the curve, they have still not reached the peak of infection, so control measures should be applied still at this time, because the number of infection will be higher in the coming days compared to past weeks.

## Data Availability

The data that support the findings of this study are openly available at https://github.com/UgoAvila/COVID-19.Mexico

## Acknowledgments

Ugo Avila Ponce de León is a doctoral student from Programa de Doctorado en Ciencias Biológicas of the Universidad Nacional Autónoma de México (UNAM). This paper was developed in the period of his PhD studies. Ugo Avila Ponce de León also received a fellowship (CVU: 774988) from Consejo Nacional de Ciencia y Tecnología (CONACYT). This article was supported in part by Mexican SNI under CVU 15284.

## Notes

### Competing Interest Statement

The authors have declared no competing interest.

### Funding Statement

Ugo Avila-Ponce de León received a fellowship (CVU: 774988) from Consejo Nacional de Ciencia y Tecnología (CONACYT).
This article was supported in part by Mexican Sistema Nacional de Investigadores under CVU 15284.

## References

[1] G. Zhang et al. “Analysis of clinical characteristics and laboratory findings of 95 cases of 2019 novel coronavirus pneumonia in Wuhan, China: a retrospective analysis”. Respiratory Research 21.1 (2020), pp. 1–10.

[2] Coronaviridae Study Group of the International Committee on Taxonomy of Viruses. “The species Severe acute respiratory syndrome-related coronavirus: classifying 2019-nCoV and naming it SARS-CoV-2”. Nature Microbiology 5 (2020), pp. 536–544.

[3] D. L. Urso. “Coronavirus Disease 2019 (COVID-19): A Brief Report”. Clinical Management Issues 14.1 (2020).

[4] G. Cruz-Pacheco et al. “Dispersion of a new coronavirus SARS-CoV-2 by airlines in 2020: Temporal estimates of the outbreak in Mexico.” medRxiv (2020).

[5] M. M. Alvarez, E. Gonzalez-Gonzalez, and G. Trujillo-de Santiago. “Modeling COVID-19 epidemics in an Excel spreadsheet: Democratizing the access to first-hand accurate predictions of epidemic outbreaks”. medRxiv (2020).

[6] M. A. Acuña-Zegarra et al. “The SARS-CoV-2 epidemic outbreak: a review of plausible scenarios of containment and mitigation for Mexico”. medRxiv (2020).

[7] Q. Lin et al. “A conceptual model for the coronavirus disease 2019 (COVID-19) outbreak in Wuhan, China with individual reaction and governmental action”. International Journal of Infectious Diseases 93 (2020), pp. 211–216.

[8] S. Zhao and H. Chen. “Modeling the epidemic dynamics and control of COVID-19 outbreak in China”. Quantitative Biology 8 (2020), pp. 11–19.

[9] D. Fanelli and F. Piazza. “Analysis and forecast of COVID-19 spreading in China, Italy and France”. Chaos, Solitons & Fractals 134 (2020), p. 109761.

[10] D. Caccavo. “Chinese and Italian COVID-19 outbreaks can be correctly described by a modified SIRD model”. medRxiv (2020).

[11] S. S. Nadim, I. Ghosh, and J. Chattopadhyay. “Short-term predictions and prevention strategies for COVID-2019: A model based study”. arXiv preprint arXiv:2003.08150 (2020).

[12] H. S. Rodrigues, M. T. T. Monteiro, and D. F. M. Torres. “Sensitivity analysis in a dengue epidemiological model”. Conference Papers in Science 2013 (2013), article ID 721406.

[13] B. Tang et al. “An updated estimation of the risk of transmission of the novel coronavirus (2019-nCov)”. Infectious Disease Modelling 5 (2020), pp. 248–255.

[14] COVID-19 Tablero México. URL: http://datos.covid-19.conacyt.mx/index.php.

[15] U. Avila-Ponce de León, Á. G. C. Pérez, and E. Avila-Vales. “A data driven analysis and forecast of an SEIARD epidemic model for COVID-19 in Mexico”. arXiv preprint arXiv:2004-08288 (2020).

